# Malaria prevalence after mass drug administration and indoor residual spraying in a conflict-affected setting, Democratic Republic of the Congo: a non-randomised repeated cross-sectional study, 2022–2024

**DOI:** 10.64898/2026.11.25.26368970

**Authors:** Jonathan Polonsky, Mahamadou Maiga, Arnaud Desjardins, Mouhamed Abdou Salam Mbengue, Patrick Nkemenang, Sophie Meakin, Esther Sterk, Jean-Patrick Ouamba, Yves Katuala, Herman Jakisa Uluba, Andrea Incerti, Etienne Gignoux, Iza Ciglenecki, Lise Grout

**Affiliations:** Epicentre, Paris, France; Geneva Centre of Humanitarian Studies, University of Geneva, Geneva, Switzerland; Médecins Sans Frontières, Operational Centre Geneva, Geneva, Switzerland; Programme National de Lutte contre le Paludisme, Kinshasa, Democratic Republic of the Congo; Zone de Santé d’Angumu, Angumu, Democratic Republic of the Congo

**Keywords:** Mass drug administration, Indoor residual spraying, Pyronaridine-artesunate, Dihydroartemisinin-piperaquine, Clothianidin, *Plasmodium falciparum*, Humanitarian emergency, Internally displaced persons, Holoendemic, Democratic Republic of the Congo

## Abstract

**Background:** The World Health Organization conditionally recommends mass drug administration (MDA) for short-term reductions in malaria burden, including in emergencies. We assessed the association of a time-limited MDA and indoor residual spraying (IRS) campaign with malaria prevalence and mortality up to 14 months after its last MDA round in Angumu, a malaria holoendemic and conflict-affected health zone in the Democratic Republic of the Congo.

**Methods:** Fourteen health areas were allocated non-randomly to MDA+IRS (those with the highest malaria incidence), IRS alone (those with the least MSF support for case management), or routine care (Routine). Malaria prevalence by rapid diagnostic test was measured at 0, 7, 13, 18, and 24 months from March 2022, and under-five and crude mortality at 0, 13, and 24 months. Within each survey, zones were compared by mixed-effects regression adjusted for area-level baseline values and individual displacement status. Routine areas were excluded after receiving interventions, leaving one at 24 months.

**Results:** At every later survey, the odds of malaria in the MDA+IRS zone were approximately a quarter to a third of those in the Routine zone. This difference persisted to the last survey, 14 months after the last MDA round, with prevalence of 24.4% compared with 56.4% (odds ratio [OR] 0.31, 95% CI 0.19–0.50). IRS alone was associated with lower prevalence during the campaign, but not statistically significantly afterwards. Prevalence was lower in the MDA+IRS than in the IRS zone at 18 and 24 months (ORs 0.37 and 0.47); during the campaign, the difference was not statistically significant after adjustment for baseline prevalence. Mortality differences were not statistically significant, except lower crude mortality in the MDA+IRS than the Routine zone at 13 months. In 2023, the sporozoite rate was lower in the intervention zones than the Routine zone, and facility test positivity was lowest in the MDA+IRS zone at every later survey.

**Conclusions:** In holoendemic, conflict-affected settings, malaria programmes and humanitarian organisations may consider co-timed MDA and IRS where high coverage is achievable, with prevalence monitored in repeated surveys to decide when to repeat the campaign.

## Background

Malaria caused an estimated 610,000 deaths worldwide in 2024, approximately 95% in the World Health Organization (WHO) African Region [1], with most malaria deaths occurring among children under five [1]. Among people in humanitarian crises, crowding, malnutrition, and disrupted case management increase malaria incidence and case fatality [2,3]. The Democratic Republic of the Congo (DRC) alone accounted for approximately one-eighth of global cases and deaths in 2024 [1].

Mass drug administration (MDA) is the treatment of the whole population of a defined area with a full antimalarial course at approximately the same time, regardless of symptoms or infection status [4]. WHO conditionally recommends MDA for short-term reductions in malaria burden in moderate-to-high transmission and in emergencies [5]. In a Cochrane review, MDA lowered *Plasmodium falciparum* prevalence one to three months after treatment in very-low-to-low transmission settings, but the effect was attenuated by four to six months [6]. There was no evidence of an effect in moderate-to-high transmission settings [6,7].

MDA clears infection in those treated, which reduces the human parasite reservoir, while indoor residual spraying (IRS) of insecticide on inner house walls shortens the survival of mosquitoes that rest on them, which reduces the number that survive until they become infectious. Transmission models predict that vector control, by IRS or insecticide-treated nets (ITNs), lengthens the effect of MDA [8,9]. In one review, 63% of MDA studies had less than six months of follow-up, and sustained reductions were reported mainly in low-transmission, island, or remote settings, usually in combination with vector control [10]. No evaluation in high-transmission sub-Saharan Africa had measured malaria prevalence more than six months after the last round of a time-limited MDA and IRS campaign.

Angumu health zone, in Ituri province, eastern DRC, is holoendemic for malaria [11]. Since December 2017, violence within Ituri province has created mass displacement of populations into and within Angumu [11]. A mortality survey in January 2020 found a crude mortality rate (CMR) and an under-five mortality rate (U5MR) above emergency thresholds [11]. Between 2019 and 2021, households reported malaria or anaemia as the cause of 66% to 72% of under-five deaths [11].

From September 2020 to January 2021, the Ministry of Health and Médecins Sans Frontières (MSF) delivered three MDA rounds in four health areas [11]. This 2020 MDA campaign was associated with a halving of U5MR over approximately 10 weeks of follow-up [11]. Neither how long the lower U5MR lasted nor any association with malaria infection was measured [11]. The present study measured both after a second campaign.

From late June 2022 to May 2023, the Ministry of Health and MSF delivered four MDA rounds and two IRS rounds in the health areas with the highest malaria incidence in routine data (MDA+IRS zone). Other areas received the two IRS rounds alone (IRS zone) or neither intervention (Routine zone) (Figure 1). We assessed the association of these interventions with malaria prevalence and mortality. Five population-based surveys were conducted: survey 1 before the campaign, surveys 2 and 3 during it, and surveys 4 and 5 approximately 8 and 14 months after the last MDA round. The primary outcome was malaria prevalence, compared within each survey between each intervention zone and the Routine zone. The MDA+IRS zone was also compared with the IRS zone, to estimate the difference associated with adding MDA where IRS was also delivered. Secondary outcomes were U5MR and CMR at surveys 1, 3, and 5. Entomological and routine facility surveillance were analysed as supporting evidence.

**Figure 1.**
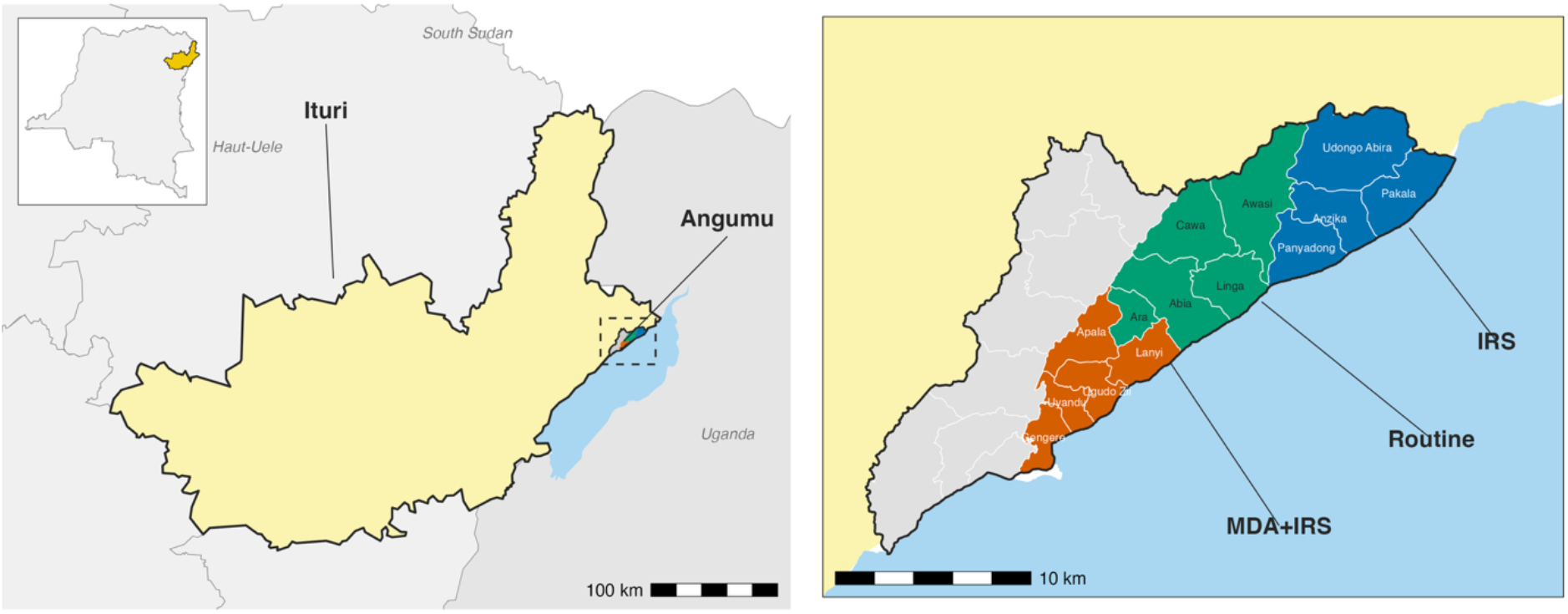
Study area and zones at survey 1, Angumu health zone, Democratic Republic of the Congo. Left: Ituri province (yellow), neighbouring provinces and countries, and Lake Albert; inset shows the position of Ituri province in the Democratic Republic of the Congo. Right: the 14 study health areas by zone at survey 1, with the eight health areas excluded for insecurity in grey. Sources: health-area and province boundaries, GRID3 COD Health Areas and Health Zones v9.0 (Center for Integrated Earth System Information, Columbia University; Ministère de la Santé Publique, Hygiène et Prévention; GRID3; CC BY); national boundary, geoBoundaries, from OpenStreetMap (ODbL); Lake Albert, © OpenStreetMap contributors (ODbL); other lakes and countries, Natural Earth. IRS, indoor residual spraying; MDA, mass drug administration.

## Methods

### Study setting and design

Angumu health zone comprises 22 health areas, eight of which were excluded from the study for insecurity [12]. The remaining 14 health areas were assigned to three intervention zones: Routine (Abia, Ara, Awasi, Cawa, and Linga), MDA+IRS (Apala, Gengere, Lanyi, Ugudo-Zii, and Uyandu), and IRS (Anzika, Pakala, Panyadong, and Udongo Abira) (Figure 1). Zone assignment was an operational decision and was not randomised: MDA+IRS was allocated to the health areas with the highest malaria incidence in routine data, and IRS alone was allocated to those health areas most lacking MSF support for routine case management. Gengere lies in a river delta on the shore of Lake Albert, where the lake spread inland between 2019 and 2021 [13], and its swamps provide breeding sites for *Anopheles*.

The 2020 MDA campaign had been delivered in health areas of both intervention zones but not in any Routine area [11]. Abia (Routine) and Gengere (MDA+IRS) had received IRS in 2021 (Additional file 1: S1 Text).

### Interventions

The MDA+IRS zone received four MDA rounds: rounds 1 and 2 with pyronaridine-artesunate in July and August 2022, and rounds 3 and 4 with dihydroartemisinin-piperaquine (DHA-piperaquine) in January and February 2023. Two IRS rounds with SumiShield 50WG (clothianidin) [14] were delivered in the MDA+IRS and IRS zones, from June to November 2022 and from February to May 2023. The campaign extended from late June 2022 to May 2023. Routine care in every zone comprised facility-based testing and treatment, and the distribution of ITNs, including a health-zone-wide distribution in December 2023 (Figure 2A). MSF also distributed ITNs in Awasi in August 2023 (MSF Operational Centre Geneva, unpublished programme data).

**Figure 2.**
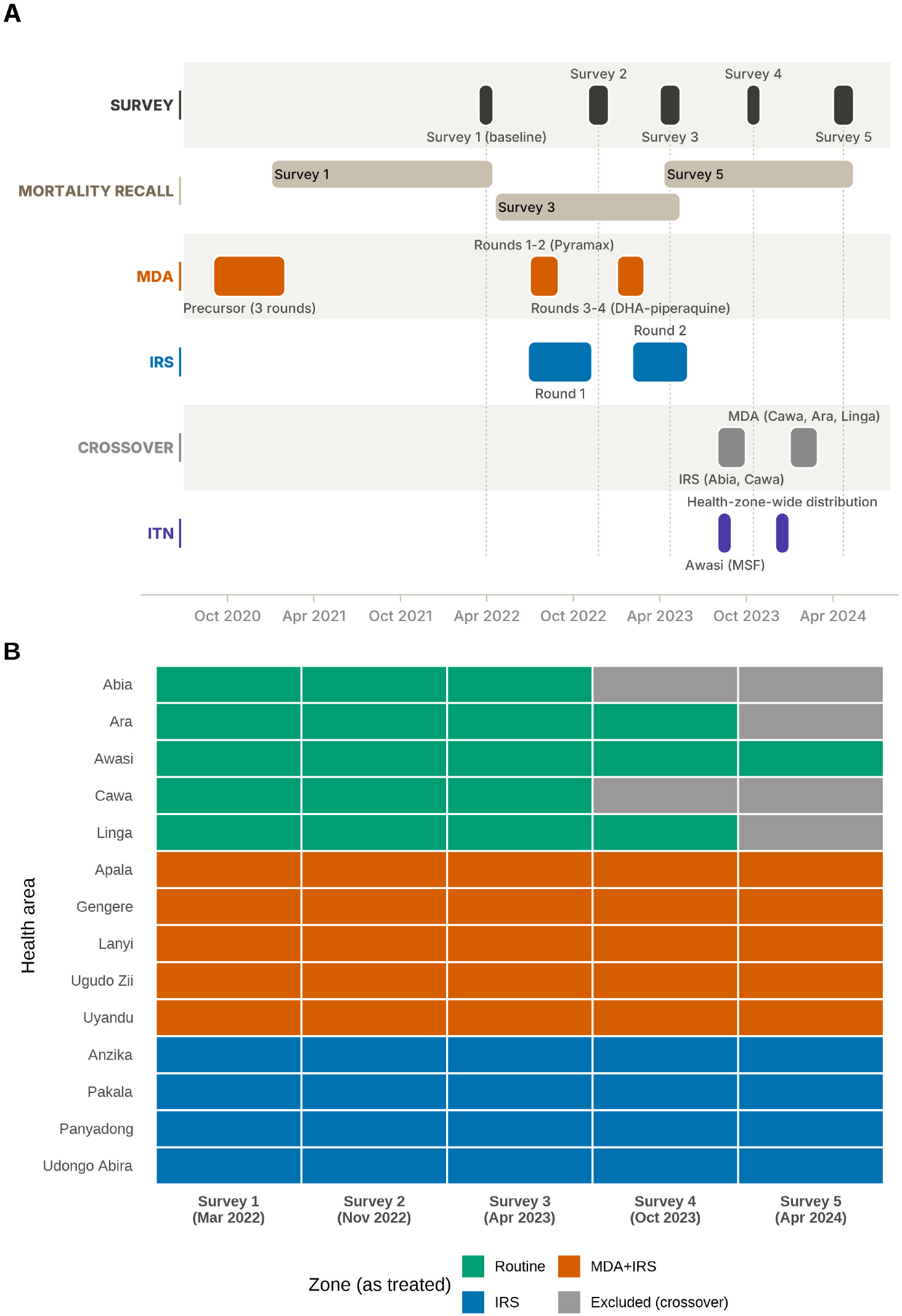
Study timeline and zone classification by survey, Angumu health zone, Democratic Republic of the Congo. (A) Timing of intervention delivery (IRS rounds, MDA rounds, the MSF ITN distribution in Awasi (August 2023), the health-zone-wide ITN distribution (December 2023), and health-area crossovers between zones), the five survey field periods, and the mortality recall periods of surveys 1, 3, and 5, January 2021 to May 2024, with the 2020 campaign for context. (B) Zone of each health area at each survey, grouped by survey-1 zone: Routine, MDA+IRS, IRS, or excluded from the zone comparison after crossover. IRS, indoor residual spraying; ITN, insecticide-treated net; MDA, mass drug administration.

Four of the five Routine health areas later received interventions (crossover areas; Figure 2B). In the primary, as-treated classification, a crossover area contributed to the Routine zone before its intervention and was excluded from the zone comparison from the following survey onwards. Abia and Cawa (IRS, August and September 2023) were excluded from survey 4 onwards, and Ara and Linga (MDA, January and February 2024) from survey 5. At survey 5, the Routine zone comprised Awasi alone, the only health area not to have received IRS or MDA up to survey 5.

### Surveys and outcomes

Five surveys were conducted between March 2022 and May 2024. Survey 1 (March to April 2022) was the baseline (Figure 2A). Months from baseline were calculated from the midpoint of each survey: 0, 7, 13, 18, and 24 at surveys 1 to 5. Each cluster comprised one rooftop, selected at random from a frame of rooftops geocoded on a satellite image dated February 2022, and its four nearest neighbours. Sample size was calculated separately for each survey (Additional file 1: S1 Text).

All household members aged 2 months or older present at the visit were tested for malaria at surveys 1, 2, and 4, and those in a random subset of households at surveys 3 and 5. Participants were tested with a rapid diagnostic test (RDT) for histidine-rich protein 2 (HRP2; SD Bioline Ag P.f., Standard Diagnostics, Republic of Korea). Malaria prevalence was the number of RDT-positive participants divided by the number with a valid result. Displacement status was recorded for each household member as displaced or resident (Additional file 1: S1 Text).

Mortality was assessed among all eligible household members by retrospective recall at surveys 1, 3, and 5, which followed the low-transmission season (the December to February dry months). The recall period ran from 1 January 2021, 18 April 2022, and 9 April 2023 at surveys 1, 3, and 5, respectively, to the interview. At survey 1, it ended before the campaign began. At survey 3, it began 10 weeks before IRS round 1 and therefore included all four MDA rounds, IRS round 1, and most of IRS round 2. At survey 5, it began after the last MDA round, in the final weeks of IRS round 2 (Figure 2A). CMR and U5MR were the number of deaths (at all ages and among children under five, respectively) divided by their person-time, expressed as deaths per 10,000 person-days.

Coverage of MDA rounds 1 and 2 and IRS round 1 was measured at survey 2 (November and December 2022), and that of the later rounds at survey 3. Intervention coverage was the proportion who had received a round (MDA) or whose dwelling had been sprayed (IRS) (Additional file 1: S1 Text).

### Entomological surveillance

Entomological surveillance data were collected from May 2022 to December 2024 with miniature Centers for Disease Control and Prevention (CDC) light traps operated indoors (Additional file 1: S3 Figure). Entomological indicators were compared descriptively between zones, to assess whether the zones ranked in the same order on these indicators as on survey malaria prevalence. From May to October 2022, the design placed 15 traps in each zone, 10 in houses selected at random from the rooftop frame and five in purposively selected traditional houses (cob walls, thatched roofs). Each trap operated for four consecutive nights a week and was moved to a different house each week, within four-week sampling cycles. In October 2022, to maintain catches in the intervention zones after IRS, five traps were withdrawn from the Routine zone and four of them were redeployed, two to each intervention zone. *Anopheles* were identified morphologically to species complex, and tested for *Plasmodium falciparum* sporozoites by circumsporozoite enzyme-linked immunosorbent assay (ELISA) [15]. Mosquitoes for testing were selected at random within each trap-week’s collection, and where catches were large, mainly in Gengere, only part of the catch was tested. Sporozoite results were used from January 2023 only, because the 2022 ELISA results were judged unreliable [15].

The sporozoite rate was the number of sporozoite-positive *Anopheles* divided by the number with a recorded laboratory result. Estimation of the human biting rate (HBR) assumed that every *Anopheles* caught in a CDC light trap was seeking a human host. HBR was the number of *Anopheles* caught in working traps (one trap in one house for one night on which it functioned) divided by the number of occupant-nights: the people who sleep in the trapped house, summed over its working trap-nights. The estimated entomological inoculation rate (EIR) was HBR multiplied by the sporozoite rate and by 365, and was reported as infective bites per person per month (the annual rate divided by 12). Because trapping was continuous, a crossover health area was excluded from the zone comparison from the month of its first intervention, rather than from the next survey as in the survey analysis. Indicators were estimated by half-year from July 2022 to November 2024, which matches the intervals between surveys. The protocol specified 2-month periods, but most held too few sporozoite-positive mosquitoes for a stable EIR, and the 2-month series is reported as supplementary (Additional file 1: S5 Figure). Residual efficacy of IRS round 2 was assessed by monthly WHO cone bioassays with local *Anopheles* on four mud walls in Panyadong (IRS zone) and four plastic-sheeting shelters in Gengere, from April to December 2023 (Additional file 1: S1 Text). Except for the pooled sporozoite rate, each was calculated per health area and averaged over a zone’s areas with population weights (Additional file 2).

### Routine facility surveillance

We analysed the monthly test positivity rate (TPR; positive RDTs divided by tests performed) in national malaria programme (PNLP) facility reports, October 2021 to October 2024. TPR was used as an indicator of malaria infection in the population, which it reflects where testing practice, care-seeking, and RDT supply are similar between zones and stable over time. Zone TPR was the unweighted mean of its health areas (13 of the 14 had data). As in the survey analysis, a crossover area was excluded from the zone comparison from the month of its first intervention. We counted the surveys at which the same zone had the lowest TPR and the lowest survey prevalence, and at which the three zones ranked identically on TPR and on survey prevalence (Additional file 1: S1 Text).

### Statistical analysis

Crude malaria prevalence, crude mortality rates, and crude odds ratios (ORs) and rate ratios (RRs) were estimated at each survey with 95% confidence intervals (CIs). Estimates were survey-weighted, with the cluster as the only sampling stage and zone as stratum. Survey-weighted malaria prevalence was also estimated for each health area, to compare areas across zones (Additional file 1: S8 Table).

In the primary model, each participant’s RDT result was modelled by mixed-effects logistic regression over the five surveys, with a zone-by-survey interaction and random intercepts for survey-specific cluster and for household within cluster. The model was adjusted for age, sex, displacement status, and area-level malaria prevalence at baseline (Additional file 1: S1 Text). Baseline prevalence was included because zones were assigned partly by prior malaria incidence. Displacement status was included because the proportion displaced at baseline was highest in the MDA+IRS zone. The primary contrast was the within-survey OR, comparing each intervention zone with the Routine zone at the same survey. The model-standardised prevalence difference between zones, in percentage points (pp), was also estimated, because an OR lies further from 1 than the prevalence ratio when prevalence is high.

The difference-in-differences (DiD) ratio of ORs, from the zone-by-survey interaction, divided the change in the odds of malaria since survey 1 in an intervention zone by the corresponding change in the Routine zone. It therefore removed any difference between zones at survey 1 that remained after adjustment. This remaining difference, the residual survey-1 OR, was the OR between zones at survey 1 after adjustment for baseline prevalence. The within-survey OR retains it.

The MDA+IRS zone was also compared with the IRS zone within each survey to estimate the difference associated with adding MDA where IRS was also delivered.

U5MR and CMR were modelled by mixed-effects Poisson regression with a log person-time offset, adjusted for sex, displacement status, and, at surveys 3 and 5, the area-level baseline value of the same rate. CMR was modelled over surveys 1, 3, and 5 together, with the zone-by-survey interaction, adjustment for age, and the random intercepts of the prevalence model, with the DiD ratio of RRs for CMR defined in the same way. U5MR was modelled separately at each survey with a cluster random intercept only, because a pooled model did not fit reliably on the small number of under-five deaths (Additional file 1: S1 Text). Within each survey, RRs were estimated for each intervention zone compared with the Routine zone, and, at surveys 3 and 5, for MDA+IRS compared with IRS.

We conducted six sensitivity analyses of prevalence. First, the Routine comparator was held fixed at Awasi at every survey, because the Routine zone fell from five health areas to three and then one. Second, a health-area random intercept was added, because zone was assigned to only 14 health areas and intervals based on clusters may be too narrow. Third, because zone was assigned to health areas, an area-level permutation test treated each health area as one observation, without adjustment for covariates. It compared the difference in mean area prevalence between two zones with the differences obtained when the health areas of those two zones were divided into two groups of the same sizes in every possible way. Fourth, E-values were calculated for the ORs comparing the MDA+IRS zone with the Routine zone at surveys 2 to 5 and with the IRS zone at surveys 4 and 5, to assess the strength of unmeasured confounding needed to explain them [16]. Fifth, two models added net use, because it differed between zones and affects malaria risk: the proportion of RDT-tested participants in each health area who slept under a net at survey 1, and, at surveys 1, 3, and 5, whether each participant slept under a net the night before. Sixth, a post-hoc analysis treated Gengere as a stratum separate from the other four MDA+IRS health areas, because its lakeside swamps supplied most of the Anopheles caught in the MDA+IRS zone and, unlike the other four areas, its prevalence changed little after survey 1. For mortality, CMR was re-estimated with Awasi as the fixed comparator, with Gengere as a separate stratum, and, at survey 3, with person-time from MDA round 1 (11 July 2022), because the survey-3 recall period began before the campaign. U5MR and CMR were also re-estimated by survey-weighted quasi-Poisson regression with design-based variance, because the mixed models assumed Poisson variance and overdispersion was not modelled. These analyses are in Additional file 1 (S2 and S3 Tables).

The pooled sporozoite rate was given an exact binomial 95% CI, and the other entomological indicators bootstrap 95% CIs from resampling of trapping sites within health areas (Additional file 1: S1 Text). No hypothesis tests were applied to the entomological indicators. Analyses used R version 4.5.2. The study is reported following the STROBE guideline for cross-sectional studies (Additional file 3).

## Results

### Study population and intervention coverage

The five surveys enrolled 621 to 3,053 households (2,597 to 15,990 household members). After exclusion of crossover areas, the zone comparison included 516 of the 621 households (83.1%) at survey 4 and 2,267 of the 3,028 (74.9%) at survey 5 (Additional file 1: S4 and S6 Tables). In the MDA+IRS zone, MDA coverage decreased from 91.2% (95% CI 87.9–93.8) at round 1 to 84.8% (95% CI 82.9–86.5) at round 4. IRS coverage was at least 94.5% (95% CI 92.2–96.1) in the MDA+IRS and IRS zones at both rounds (Additional file 1: S7 Table).

At survey 1, before the campaign, CMR was highest in the MDA+IRS zone, whereas malaria prevalence was lowest in that zone and highest in the IRS zone. CMR was 0.84 (95% CI 0.60–1.07) deaths per 10,000 person-days in the Routine zone and 1.38 (95% CI 1.15–1.60) in the MDA+IRS zone, an RR of 1.78 (95% CI 1.25–2.56) (Table 2). The CMR difference between the IRS and Routine zones, and both U5MR differences from the Routine zone, were not statistically significant. Malaria prevalence was 36.1% (95% CI 32.9–39.3) in the Routine zone, 31.4% (95% CI 28.4–34.3) in the MDA+IRS zone, and 46.8% (95% CI 43.8–49.9) in the IRS zone (Table 1). Before adjustment for baseline prevalence, the OR comparing the MDA+IRS zone with the IRS zone was 0.43 (95% CI 0.33–0.55) (Additional file 1: S1 Text). The proportion of participants who were displaced was 18% in the Routine zone, 33% in the MDA+IRS zone, and 20% in the IRS zone (Additional file 1: S1 Table).

**Table 1.** Malaria prevalence by intervention zone and survey, Angumu health zone, Democratic Republic of the Congo, 2022–2024.

| Zone | Survey | Months from baseline | Season | N (participants) | Crude malaria prevalence (95% CI) | Crude OR (95% CI) | OR (95% CI) | p-value | Prevalence difference, pp (95% CI) | DiD ratio of ORs (95% CI) |
|---|---|---|---|---|---|---|---|---|---|---|
| Routine | 1 | 0 | Low transmission | 2259 | 36.1 (32.9-39.3) | Reference | Reference | – | Reference | – |
| MDA+IRS | 1 | 0 | Low transmission | 2940 | 31.4 (28.4-34.3) | 0.81 (0.67-0.98) | 0.75 (0.59-0.97) | – | – | – |
| IRS | 1 | 0 | Low transmission | 2240 | 46.8 (43.8-49.9) | 1.56 (1.30-1.87) | 1.76 (1.35-2.28) | – | – | – |
| Routine | 2 | 7 | High transmission | 885 | 62.5 (56.4-68.5) | Reference | Reference | Reference | Reference | Reference |
| MDA+IRS | 2 | 7 | High transmission | 874 | 39.2 (33.3-45.1) | 0.39 (0.27-0.55) | 0.35 (0.23-0.54) | <0.001 | -25.3 (-35.2 to -15.4) | 0.41 (0.27-0.63) |
| IRS | 2 | 7 | High transmission | 845 | 54.7 (47.5-62.0) | 0.73 (0.49-1.07) | 0.50 (0.32-0.80) | 0.002 | -16.6 (-27.5 to -5.7) | 0.41 (0.26-0.65) |
| Routine | 3 | 13 | Low transmission | 1450 | 53.2 (48.4-58.0) | Reference | Reference | Reference | Reference | Reference |
| MDA+IRS | 3 | 13 | Low transmission | 934 | 27.0 (23.0-31.0) | 0.33 (0.25-0.43) | 0.27 (0.20-0.37) | <0.001 | -30.7 (-37.5 to -23.9) | 0.31 (0.22-0.44) |
| IRS | 3 | 13 | Low transmission | 952 | 41.4 (37.2-45.6) | 0.62 (0.48-0.81) | 0.36 (0.26-0.50) | <0.001 | -24.4 (-31.8 to -17.0) | 0.30 (0.21-0.42) |
| Routine | 4 | 18 | High transmission | 359 | 59.6 (52.4-66.8) | Reference | Reference | Reference | Reference | Reference |
| MDA+IRS | 4 | 18 | High transmission | 836 | 31.9 (24.8-39.0) | 0.32 (0.20-0.49) | 0.24 (0.13-0.42) | <0.001 | -34.3 (-47.0 to -21.7) | 0.27 (0.16-0.47) |
| IRS | 4 | 18 | High transmission | 736 | 57.1 (50.6-63.6) | 0.90 (0.61-1.35) | 0.64 (0.36-1.14) | 0.147 | -10.7 (-24.3 to 2.8) | 0.52 (0.30-0.91) |
| Routine | 5 | 24 | Low transmission | 286 | 56.4 (49.2-63.6) | Reference | Reference | Reference | Reference | Reference |
| MDA+IRS | 5 | 24 | Low transmission | 875 | 24.4 (20.5-28.3) | 0.25 (0.17-0.36) | 0.31 (0.19-0.50) | <0.001 | -26.8 (-38.1 to -15.4) | 0.36 (0.22-0.58) |
| IRS | 5 | 24 | Low transmission | 942 | 46.9 (42.2-51.6) | 0.68 (0.48-0.97) | 0.66 (0.41-1.04) | 0.079 | -10.5 (-21.9 to 1.0) | 0.54 (0.33-0.86) |

Zones as treated; Routine zone: Abia, Ara, Awasi, Cawa, and Linga at surveys 1 to 3; Ara, Awasi, and Linga at survey 4; Awasi at survey 5. N: participants with a valid RDT result. Prevalence and crude OR: survey-weighted, unadjusted. OR: mixed-effects logistic model (Methods); at survey 1, without the baseline covariate. Prevalence difference: model-standardised (pp, zone minus Routine). DiD ratio of ORs, difference-in-differences ratio of ORs versus survey 1 (Methods). CI, confidence interval; IRS, indoor residual spraying; MDA, mass drug administration; OR, odds ratio; pp, percentage points; RDT, rapid diagnostic test.

**Table 2.**
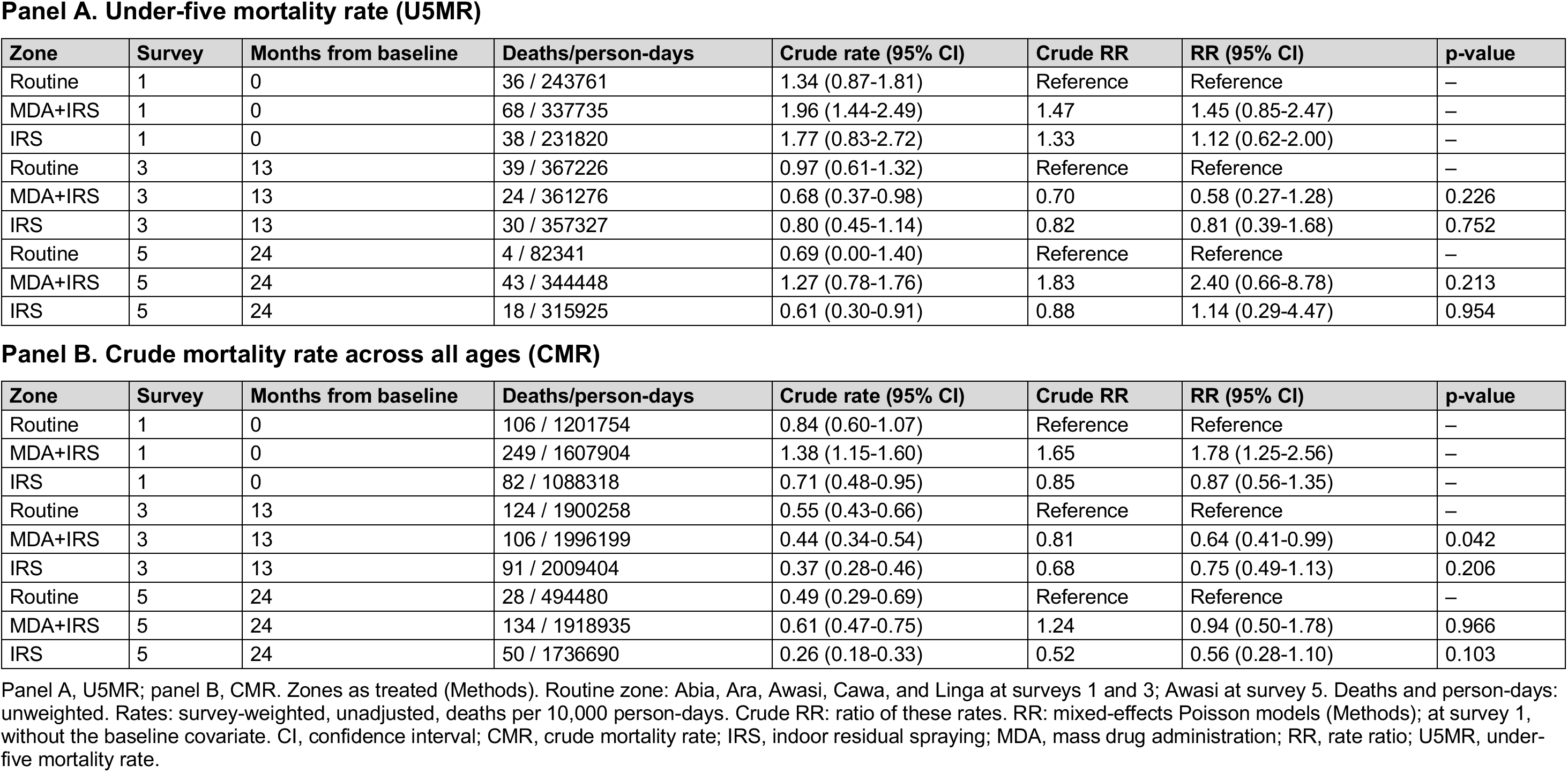
Under-five mortality rate (A) and crude mortality rate (B) by intervention zone and survey, Angumu health zone, Democratic Republic of the Congo, 2022–2024 (deaths per 10,000 person-days).

| Panel A. Under-five mortality rate (U5MR) |  |  |  |  |  |  |  |
| Zone | Survey | Months from baseline | Deaths/person-days | Crude rate (95% CI) | Crude RR | RR (95% CI) | p-value |
|---|---|---|---|---|---|---|---|
| Routine | 1 | 0 | 36 / 243761 | 1.34 (0.87-1.81) | Reference | Reference | – |
| MDA+IRS | 1 | 0 | 68 / 337735 | 1.96 (1.44-2.49) | 1.47 | 1.45 (0.85-2.47) | – |
| IRS | 1 | 0 | 38 / 231820 | 1.77 (0.83-2.72) | 1.33 | 1.12 (0.62-2.00) | – |
| Routine | 3 | 13 | 39 / 367226 | 0.97 (0.61-1.32) | Reference | Reference | – |
| MDA+IRS | 3 | 13 | 24 / 361276 | 0.68 (0.37-0.98) | 0.70 | 0.58 (0.27-1.28) | 0.226 |
| IRS | 3 | 13 | 30 / 357327 | 0.80 (0.45-1.14) | 0.82 | 0.81 (0.39-1.68) | 0.752 |
| Routine | 5 | 24 | 4 / 82341 | 0.69 (0.00-1.40) | Reference | Reference | – |
| MDA+IRS | 5 | 24 | 43 / 344448 | 1.27 (0.78-1.76) | 1.83 | 2.40 (0.66-8.78) | 0.213 |
| IRS | 5 | 24 | 18 / 315925 | 0.61 (0.30-0.91) | 0.88 | 1.14 (0.29-4.47) | 0.954 |
| Panel B. Crude mortality rate across all ages (CMR) |  |  |  |  |  |  |  |
| Zone | Survey | Months from baseline | Deaths/person-days | Crude rate (95% CI) | Crude RR | RR (95% CI) | p-value |
| Routine | 1 | 0 | 106 / 1201754 | 0.84 (0.60-1.07) | Reference | Reference | – |
| MDA+IRS | 1 | 0 | 249 / 1607904 | 1.38 (1.15-1.60) | 1.65 | 1.78 (1.25-2.56) | – |
| IRS | 1 | 0 | 82 / 1088318 | 0.71 (0.48-0.95) | 0.85 | 0.87 (0.56-1.35) | – |
| Routine | 3 | 13 | 124 / 1900258 | 0.55 (0.43-0.66) | Reference | Reference | – |
| MDA+IRS | 3 | 13 | 106 / 1996199 | 0.44 (0.34-0.54) | 0.81 | 0.64 (0.41-0.99) | 0.042 |
| IRS | 3 | 13 | 91 / 2009404 | 0.37 (0.28-0.46) | 0.68 | 0.75 (0.49-1.13) | 0.206 |
| Routine | 5 | 24 | 28 / 494480 | 0.49 (0.29-0.69) | Reference | Reference | – |
| MDA+IRS | 5 | 24 | 134 / 1918935 | 0.61 (0.47-0.75) | 1.24 | 0.94 (0.50-1.78) | 0.966 |
| IRS | 5 | 24 | 50 / 1736690 | 0.26 (0.18-0.33) | 0.52 | 0.56 (0.28-1.10) | 0.103 |

Panel A, U5MR; panel B, CMR. Zones as treated (Methods). Routine zone: Abia, Ara, Awasi, Cawa, and Linga at surveys 1 and 3; Awasi at survey 5. Deaths and person-days: unweighted. Rates: survey-weighted, unadjusted, deaths per 10,000 person-days. Crude RR: ratio of these rates. RR: mixed-effects Poisson models (Methods); at survey 1, without the baseline covariate. CI, confidence interval; CMR, crude mortality rate; IRS, indoor residual spraying; MDA, mass drug administration; RR, rate ratio; U5MR, under-five mortality rate.

### Malaria prevalence

Malaria prevalence in the Routine zone was higher at every later survey than at survey 1, whereas in the MDA+IRS zone it was below its survey-1 level at surveys 3 and 5 (Table 1; Figures 3 and 4). In the Routine zone, prevalence was 53.2% (95% CI 48.4–58.0) at survey 3 and 56.4% (95% CI 49.2–63.6) at survey 5, above Awasi’s survey-1 level of 44.1% (Additional file 1: S2 Table, panel B). In the MDA+IRS zone, prevalence was 27.0% (95% CI 23.0–31.0) at survey 3 and 24.4% (95% CI 20.5–28.3) at survey 5.

**Figure 3.**
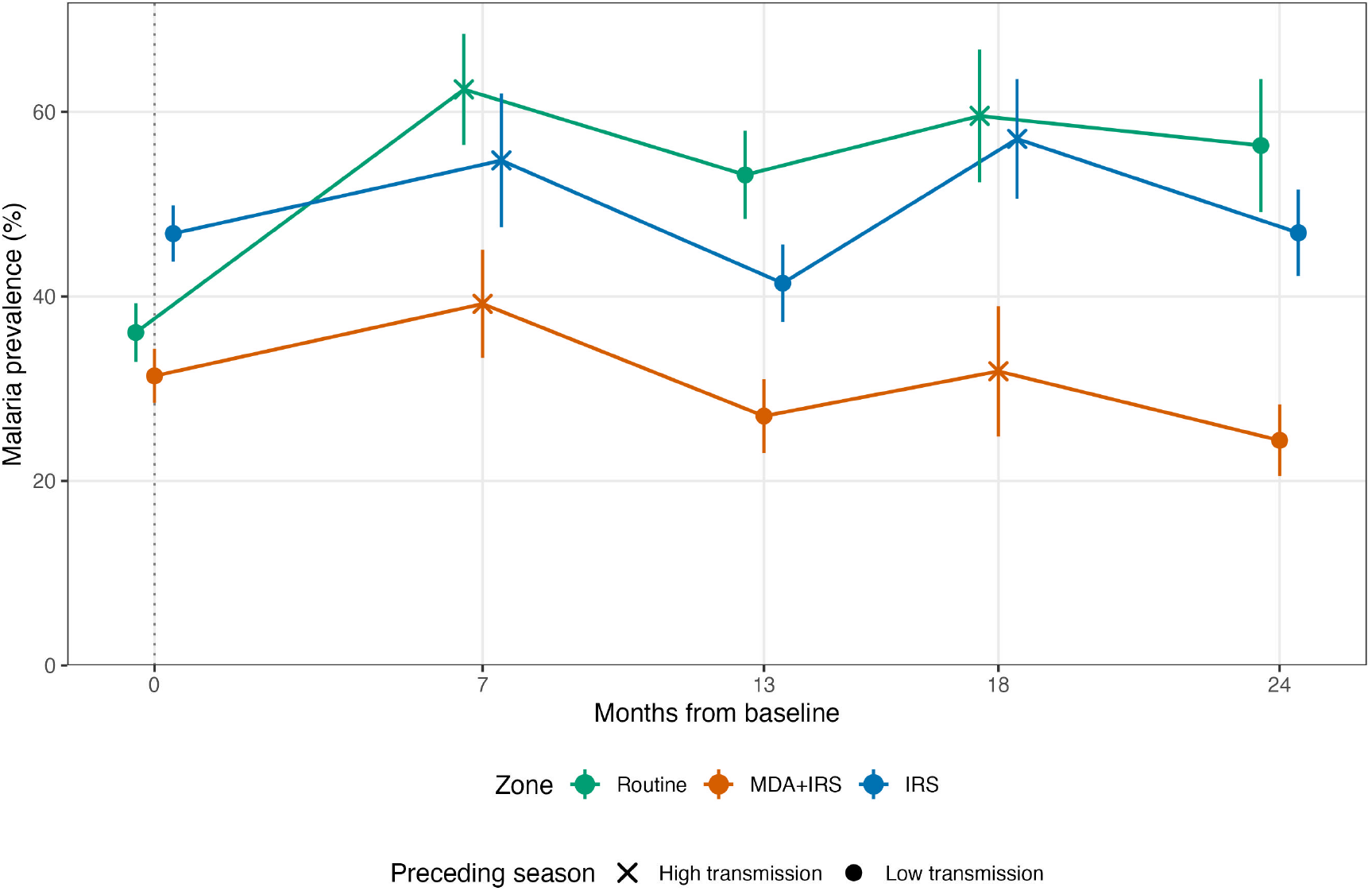
Observed malaria prevalence (%) with 95% confidence intervals by intervention zone and months from baseline. Circles mark surveys that followed the low-transmission (dry) months and crosses those that followed the high-transmission (rainy) months. The dotted vertical line marks survey 1, before the interventions. Points are offset horizontally by zone for legibility. The Routine zone comprised five health areas at surveys 1 to 3, three at survey 4, and Awasi alone at survey 5. IRS, indoor residual spraying; MDA, mass drug administration.

**Figure 4.**
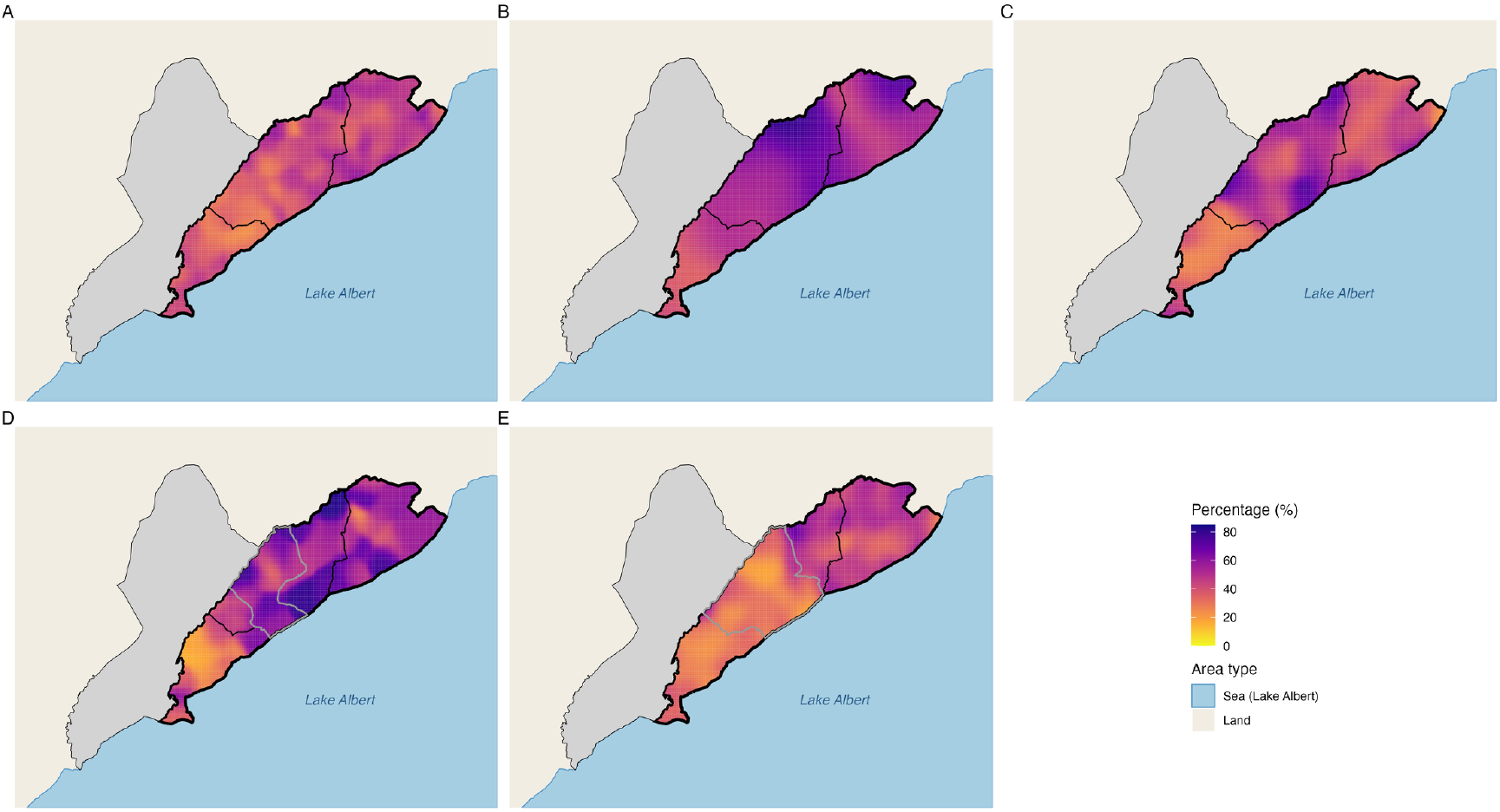
Smoothed malaria prevalence (%) by survey, Angumu health zone, Democratic Republic of the Congo, 2022–2024. (A) Survey 1, March to April 2022; (B) survey 2, November to December 2022; (C) survey 3, April to May 2023; (D) survey 4, October 2023; (E) survey 5, April to May 2024. Grey outlines in panels D and E mark health areas excluded from the zone comparison after crossover.

At each of surveys 2 to 5, survey-weighted malaria prevalence was lower in every MDA+IRS health area than in every Routine health area (Additional file 1: S4 Figure). At survey 1, the ranges overlapped: 20.4% to 42.0% in the MDA+IRS areas and 28.8% to 44.1% in the Routine areas (Additional file 1: S8 Table). In the area-level permutation test, the p-value for the MDA+IRS zone compared with the Routine zone was 0.008 at surveys 2 and 3 and 0.018 at survey 4 (Additional file 1: S2 Table, panel D). At survey 5, it was p=0.167, the smallest value attainable with six health areas.

The odds of malaria in the MDA+IRS zone were a quarter to a third of those in the Routine zone at each survey after the campaign began (OR 0.24 to 0.35; p<0.001 at each survey; Table 1), including 0.31 (95% CI 0.19–0.50) at survey 5, 14 months after the last MDA round. Model-standardised prevalence was 25.3 to 34.3 pp lower than in the Routine zone. The difference was larger than at survey 1, when the residual survey-1 OR was 0.87 (95% CI 0.67–1.11), and the DiD ratio of ORs was 0.27 to 0.41, with its 95% CI excluding 1 at every survey (Additional file 1: S2 Table, panel A).

In the IRS zone, the odds of malaria were about half those in the Routine zone at survey 2 (OR 0.50, 95% CI 0.32–0.80) and a third at survey 3 (OR 0.36, 95% CI 0.26–0.50), during the campaign, and the differences were not statistically significant after the campaign (OR 0.64, 95% CI 0.36–1.14, at survey 4; 0.66, 95% CI 0.41–1.04, at survey 5; Table 1). In the area-level permutation test, the difference was not statistically significant at any survey (Additional file 1: S2 Table, panel D). The residual survey-1 OR was 1.22 (95% CI 0.93–1.61), and the DiD ratio of ORs was 0.52 (95% CI 0.30–0.91) at survey 4 and 0.54 (95% CI 0.33–0.86) at survey 5, with its 95% CI excluding 1 at every survey (Additional file 1: S2 Table, panel A).

With Awasi as the fixed comparator or a health-area random intercept, the MDA+IRS ORs (0.20 to 0.35) were statistically significant at surveys 2 to 5, and the IRS ORs at surveys 2 and 3 only (Additional file 1: S2 Table, panels B and C). Adjustment for area-level or individual net use changed the MDA+IRS ORs by 0.02 or less and the IRS ORs by 0.10 or less, and did not change which contrasts were statistically significant (Additional file 1: S2 Table, panel G; S11 Table). With Gengere as a separate stratum, the MDA+IRS ORs at surveys 2 to 5 were 0.19 to 0.37 in the other four health areas and 0.31 to 0.49 in Gengere, and the difference between the two strata was not statistically significant (joint test p=0.057; Additional file 1: S2 Table, panel H).

After the campaign, the odds of malaria in the MDA+IRS zone were less than half those in the IRS zone (OR 0.37, 95% CI 0.23–0.60, at survey 4; 0.47, 95% CI 0.33–0.68, at survey 5; p<0.001 at each), compared with a residual survey-1 OR of 0.71 (95% CI 0.53–0.94). At surveys 2 and 3, during the campaign, the difference between the two zones was not statistically significant in the primary model (Additional file 1: S2 Table, panel E). In the area-level permutation test, mean area prevalence in the MDA+IRS zone was 15.4 and 16.7 percentage points below that in the IRS zone at surveys 2 and 3 (p=0.024 at each), close to the 15.1-point difference at survey 1, and 28.3 and 22.7 points below at surveys 4 and 5 (p=0.016 and 0.024) (Additional file 1: S2 Table, panel D). The E-values for the MDA+IRS OR were 2.75 to 3.52 compared with the Routine zone at surveys 2 to 5 (2.05 to 2.68 for the 95% CI limits closest to 1) and 2.67 and 2.28 compared with the IRS zone at surveys 4 and 5 (1.91 and 1.73), against 1.36 and 1.66 for the residual survey-1 ORs (Additional file 1: S2 Table, panel F).

### Mortality

At surveys 3 and 5, U5MR was 0.61 to 1.27 and CMR 0.26 to 0.61 deaths per 10,000 person-days across the three zones (Table 2; Additional file 1: S2 Figure). U5MR in the MDA+IRS zone was 0.68 (95% CI 0.37–0.98) at survey 3 and 1.27 (95% CI 0.78–1.76) at survey 5, the highest of the three zones at that survey.

The differences in U5MR between each intervention zone and the Routine zone were not statistically significant at survey 3 or survey 5 (Table 2). At survey 3, the RR was 0.58 (95% CI 0.27–1.28) in the MDA+IRS zone and 0.81 (95% CI 0.39–1.68) in the IRS zone. Only four under-five deaths were recorded in the Routine zone at survey 5, when the RR was 2.40 (95% CI 0.66–8.78) in the MDA+IRS zone and 1.14 (95% CI 0.29–4.47) in the IRS zone.

CMR in the MDA+IRS zone was approximately two-thirds of that in the Routine zone over the survey-3 recall period (RR 0.64, 95% CI 0.41–0.99; p=0.042; Table 2), and the difference was not statistically significant over the survey-5 recall period (RR 0.94, 95% CI 0.50–1.78). The DiD ratio of RRs was 0.46 (95% CI 0.29–0.73) at survey 3 and 0.68 (95% CI 0.36–1.27) at survey 5 (Additional file 1: S3 Table, panel A). In the IRS zone, the CMR differences from the Routine zone were not statistically significant at either survey. The differences in U5MR and CMR between the MDA+IRS and IRS zones were not statistically significant at survey 3 or survey 5 (Additional file 1: S3 Table, panel D). In the design-based analysis, every CMR contrast and every survey-3 U5MR contrast was on the same side of 1 as in the primary analysis (Additional file 1: S3 Table, panel E). No mortality contrast changed statistical significance. With Gengere as a separate stratum, the survey-3 CMR RR was 0.69 (95% CI 0.41–1.16) in the other four MDA+IRS health areas and 0.60 (95% CI 0.27–1.32) in Gengere, and the difference between the strata in the change from survey 1 was not statistically significant over surveys 3 and 5 (joint test p=0.157; Additional file 1: S3 Table, panel F). U5MR in Gengere was based on 3 and 8 deaths at surveys 3 and 5.

### Entomological indicators

In 2023, the sporozoite rate was 2.3% to 4.4% in the intervention zones, below the 10.8% (95% CI 8.6–13.3) and 12.0% (95% CI 9.1–15.4) in the Routine zone (Table 3; Figure 5C). From July to November 2024, it was lowest in the MDA+IRS zone, at 0.3% (95% CI 0.1–0.7; 7 of 2,062), based almost entirely on mosquitoes from Gengere (96% of those tested) (Additional file 2: S5 Table). In the IRS zone, the sporozoite rate rose in each half-year from January 2023.

**Table 3.** Entomological indicators by intervention zone and half-year, Angumu health zone, Democratic Republic of the Congo, July 2022 to November 2024.

| Period | Months after IRS round 2 | Zone | Working trap-nights | Anopheles in working traps | Mosquitoes tested (positive) | Sporozoite rate % (95% CI) | HBR per occupant-night (95% CI) | EIR per person per month (95% CI) |
|---|---|---|---|---|---|---|---|---|
| Jul 2022 to Dec 2022 | – | Routine | 1,709 | 3,344 | – | – | 0.35 (0.28-0.47) | – |
| Jul 2022 to Dec 2022 | – | MDA+IRS | 1,650 | 5,280 | – | – | 0.89 (0.29-1.65) | – |
| Jul 2022 to Dec 2022 | – | IRS | 1,302 | 740 | – | – | 0.12 (0.08-0.15) | – |
| Jan 2023 to Jun 2023 | during/at end | Routine | 949 | 769 | 703 (76) | 10.8 (8.6-13.3) | 0.15 (0.10-0.20) | 0.51 (0.31-0.78) |
| Jan 2023 to Jun 2023 | during/at end | MDA+IRS | 1,637 | 866 | 700 (26) | 3.7 (2.4-5.4) | 0.13 (0.06-0.17) | 0.15 (0.031-0.27) |
| Jan 2023 to Jun 2023 | during/at end | IRS | 1,649 | 178 | 173 (4) | 2.3 (0.6-5.8) | 0.02 (0.01-0.03) | 0.012 (0-0.026) |
| Jul 2023 to Dec 2023 | 1-7 | Routine | 405 | 539 | 441 (53) | 12.0 (9.1-15.4) | 0.19 (0.12-0.29) | 0.63 (0.35-1.5) |
| Jul 2023 to Dec 2023 | 1-7 | MDA+IRS | 1,481 | 3,422 | 1,029 (25) | 2.4 (1.6-3.6) | 0.82 (0.12-1.50) | 0.54 (0.050-1.0) |
| Jul 2023 to Dec 2023 | 1-7 | IRS | 1,452 | 523 | 520 (23) | 4.4 (2.8-6.6) | 0.08 (0.04-0.13) | 0.10 (0.030-0.19) |
| Jan 2024 to Jun 2024 | 7-13 | Routine | 240 | 77 | 77 (3) | 3.9 (0.8-11.0) | 0.08 (0.03-0.18) | 0.089 (0-0.15) |
| Jan 2024 to Jun 2024 | 7-13 | MDA+IRS | 1,459 | 1,997 | 1,454 (26) | 1.8 (1.2-2.6) | 0.45 (0.08-0.89) | 0.22 (0.023-0.46) |
| Jan 2024 to Jun 2024 | 7-13 | IRS | 1,455 | 221 | 227 (12) | 5.3 (2.8-9.1) | 0.03 (0.02-0.05) | 0.049 (0.0059-0.086) |
| Jul 2024 to Nov 2024 | 13-18 | Routine | 416 | 234 | 230 (20) | 8.7 (5.4-13.1) | 0.16 (0.08-0.24) | 0.43 (0.14-0.68) |
| Jul 2024 to Nov 2024 | 13-18 | MDA+IRS | 1,124 | 2,060 | 2,062 (7) | 0.3 (0.1-0.7) | 0.50 (0.10-1.23) | 0.052 (0.0075-0.11) |
| Jul 2024 to Nov 2024 | 13-18 | IRS | 663 | 104 | 107 (8) | 7.5 (3.3-14.2) | 0.04 (0.02-0.07) | 0.083 (0.029-0.16) |

Zones as treated, by month (Methods); Routine zone: Awasi alone from January 2024. Months after IRS round 2: from its end (May 2023) to the start and end of the period. Sporozoite rate: pooled, exact binomial 95% CI. HBR: *Anopheles* caught in working traps per occupant-night. EIR: infective bites per person per month. Values per working trap-night are in Additional file 2, and the 2-month series in Additional file 1. HBR and EIR: population-weighted means of health-area values, bootstrap percentile 95% CIs. Dashes: not estimated (Methods). CI, confidence interval; EIR, entomological inoculation rate; HBR, human biting rate; IRS, indoor residual spraying; MDA, mass drug administration.

**Figure 5.**
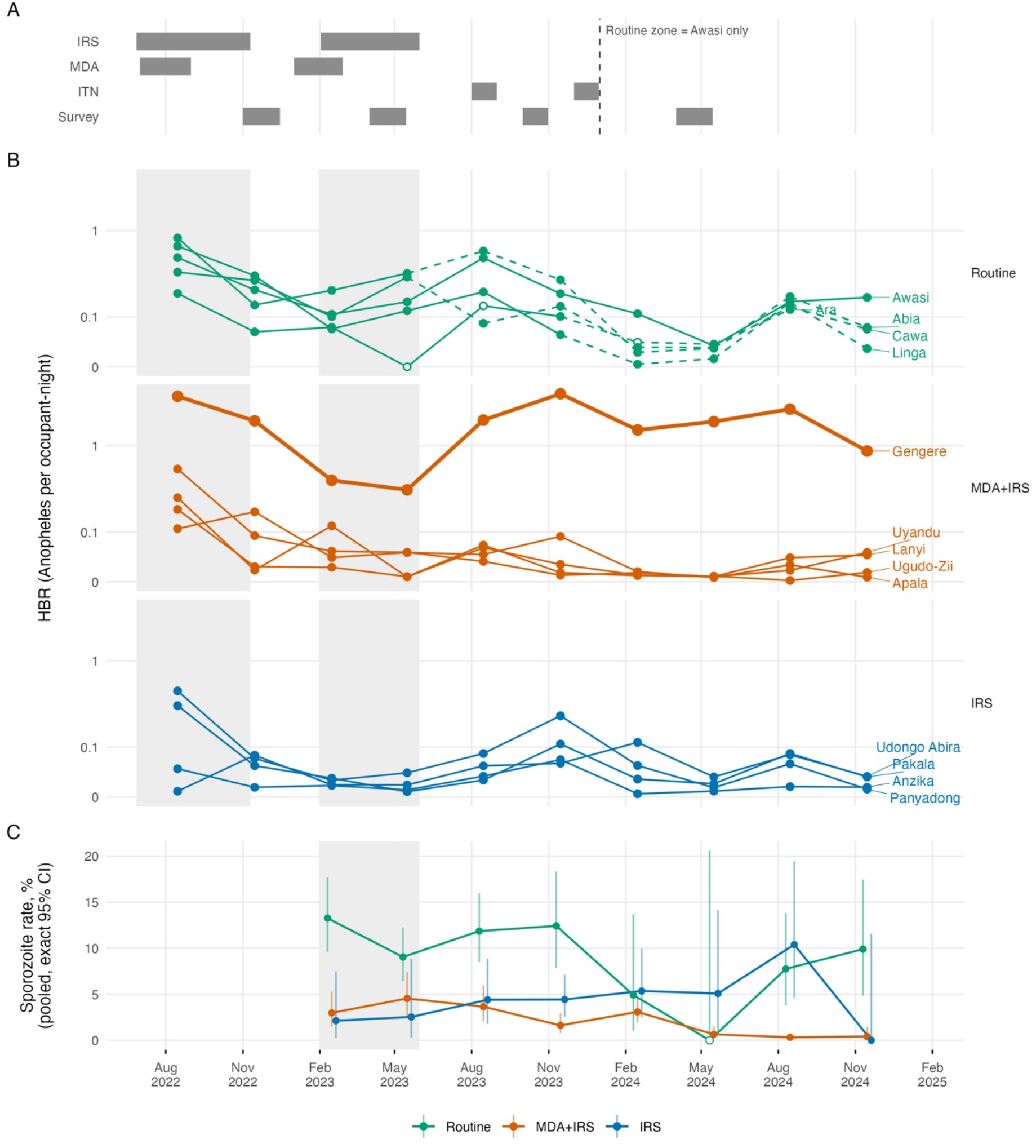
HBR by health area and sporozoite rate by zone, by quarter, Angumu health zone, Democratic Republic of the Congo, 2022–2024. (A) Timing of IRS, MDA, the ITN distributions (August 2023, Awasi only; December 2023, health-zone-wide), and surveys 2 to 5; dashed vertical line, Routine zone restricted to Awasi from January 2024. (B) HBR (*Anopheles* per occupant-night), one panel per zone at survey 1 and one line per health area, on a pseudo-logarithmic axis; Gengere, thick line; dashed segments, health area after leaving the Routine zone; open symbols, fewer than 30 working trap-nights. (C) Pooled sporozoite rate by zone (MDA+IRS including Gengere), exact binomial 95% CIs, quarters with at least 10 mosquitoes tested; open symbols, fewer than 30 tested. Grey bands: IRS rounds. CI, confidence interval; HBR, human biting rate; IRS, indoor residual spraying; ITN, insecticide-treated net; MDA, mass drug administration.

Estimated HBR was lowest in the IRS zone in each of the five half-years from July 2022, and highest in the MDA+IRS zone in four of them (Table 3). In each half-year, 75% to 98% of the *Anopheles* caught in the MDA+IRS zone were caught in Gengere. HBR was lower from July to November 2024 than from July to December 2022 in 9 of the 10 health areas trapped in both periods, and unchanged in Udongo Abira (Figure 5B; Additional file 2: S5 Table). Awasi received neither IRS nor MDA, but received an MSF ITN distribution in August 2023 and the health-zone-wide distribution in December 2023. In Awasi, HBR from July to November 2024 was 0.43 times (95% CI 0.22–0.77) its level from July to December 2022.

Estimated EIR was lower in both intervention zones than in the Routine zone from January to June 2023 (Table 3). It was 0.51 (95% CI 0.31–0.78) infective bites per person per month in the Routine zone, 0.15 (95% CI 0.031–0.27) in the MDA+IRS zone, and 0.012 (95% CI 0–0.026) in the IRS zone. From July to December 2023, when the catch in Gengere was approximately five times that of the first half, EIR was 0.54 (95% CI 0.050–1.0) in the MDA+IRS zone and 0.63 (95% CI 0.35–1.5) in the Routine zone. EIR was lowest in the IRS zone in each half-year from January 2023 to June 2024, and lowest in the MDA+IRS zone from July to November 2024. In WHO cone bioassays with local Anopheles after IRS round 2, Abbott-corrected 72-hour mortality on plastic sheeting in Gengere remained at or above 80% up to 245 days after spraying, whereas on mud walls in Panyadong (IRS zone) it was below 80% in the valid tests from day 179, before survey 4 (Additional file 1: S12 Table).

### Routine facility surveillance

Facility TPR and survey malaria prevalence were both lowest in the MDA+IRS zone at each of surveys 2 to 5 (Figure 6). The three zones ranked in the same order on both measures at surveys 3 and 5. At survey 1, TPR was lowest in the IRS zone, whereas survey malaria prevalence was lowest in the MDA+IRS zone. Before the campaign, from October 2021 to May 2022, monthly TPR in the MDA+IRS zone was 0.1 to 16.9 pp below that in the Routine zone, and it fell from 79.3% to 48.4% between June and August 2022, while TPR in the other two zones changed little (Additional file 1: S1 Figure, S10 Table). TPR in the MDA+IRS zone was above that in the Routine zone in four of the five months from July to November 2023 and below it in seven of the nine months from February to October 2024, when the Routine zone comprised Awasi alone.

**Figure 6.**
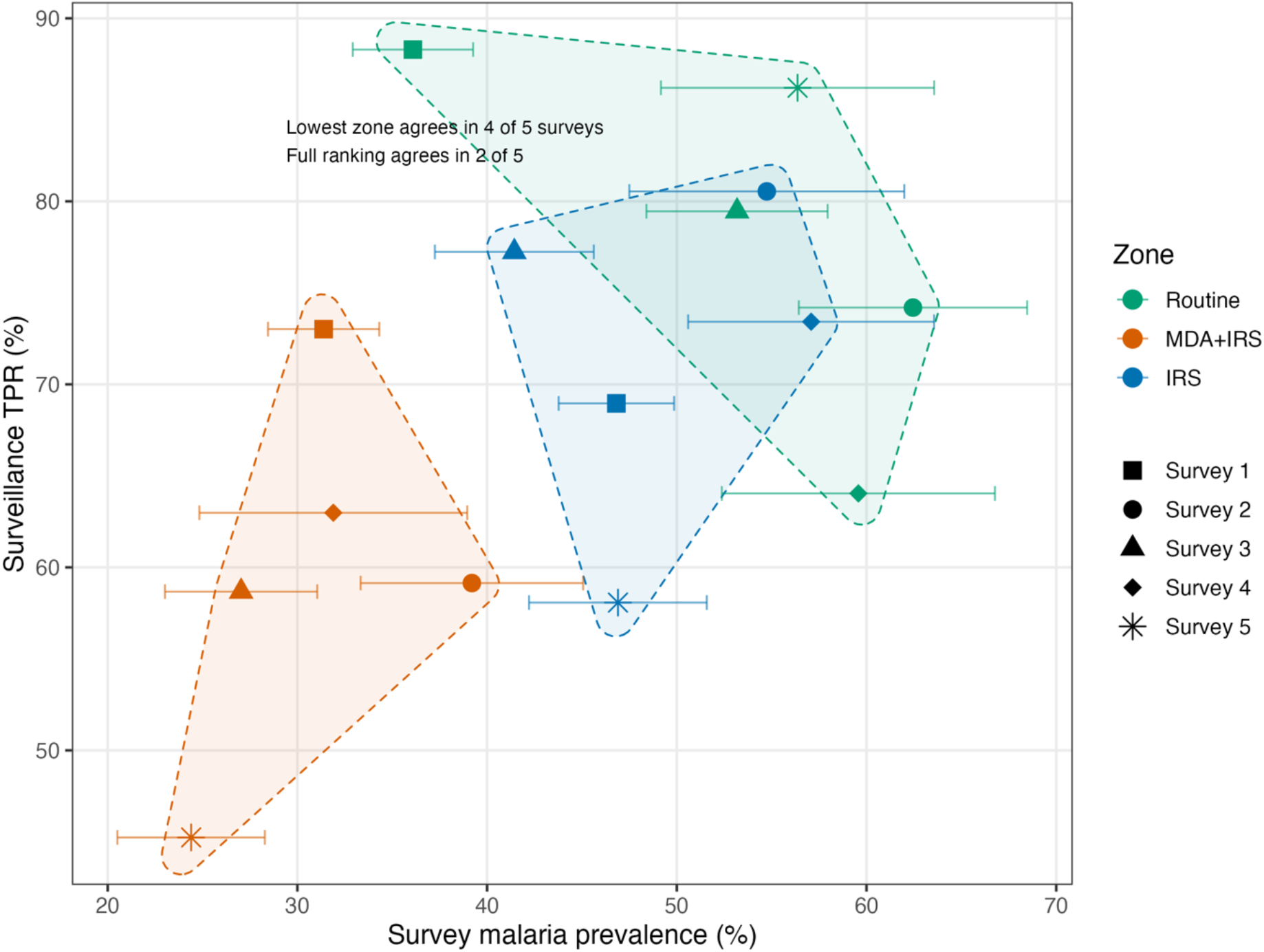
Facility TPR and survey malaria prevalence by zone and survey, Angumu health zone, 2022–2024. Each point is one zone at one survey: survey-weighted malaria prevalence with its 95% CI (horizontal bars) against facility TPR in the calendar month assigned to the survey. Colour: zone; symbol: survey. Shaded outlines enclose the points of each zone. The annotation gives the number of surveys at which the two measures agreed on the lowest zone and on the full ranking of zones. Kendall’s W: 0.67 (Additional file 1: S1 Text). Routine zone at survey 5: Awasi alone. CI, confidence interval; IRS, indoor residual spraying; MDA, mass drug administration; TPR, test positivity rate.

## Discussion

### Principal findings

Malaria prevalence was lower in the MDA+IRS zone than in the Routine zone at every survey after baseline, a difference persisting to the last survey, 14 months after the last MDA round. IRS alone was associated with lower prevalence during the campaign in the primary model, but not when health areas were compared as units without adjustment for baseline prevalence, and the association weakened after spraying ended. Prevalence was lower in the MDA+IRS zone than in the IRS zone at surveys 4 and 5, the difference associated with adding MDA where IRS was also delivered. CMR was lower in the MDA+IRS zone than in the Routine zone at survey 3 only, and the differences in U5MR between zones were not statistically significant.

### Persistence of lower prevalence with MDA+IRS

Lower malaria prevalence in the MDA+IRS zone lasted far longer than the protection that the drugs give. The effect of MDA wanes within one to three months [5], and DHA-piperaquine, used in MDA rounds 3 and 4, protects against reinfection for several weeks [17]. Survey 3, approximately two months after the last MDA round, may partly reflect that protection, but surveys 4 and 5 were beyond it. One explanation consistent with this pattern is that MDA cleared the parasite reservoir while IRS shortened mosquito survival, which reduced the number that survived until they became infectious. After spraying ended, the reservoir would then have refilled slowly, because fewer infected people remained to infect mosquitoes than in the zones without MDA. In a transmission model, co-timed IRS and MDA had approximately twice the impact of sequential delivery, because IRS protected the cleared reservoir from reinfection [8]. Consensus modelling in low-transmission settings predicted that the effect of MDA would be temporary unless vector control was scaled up [18]. However, prevalence in the MDA+IRS zone was still lower at survey 5, approximately 11 months after spraying ended.

Chance at the level of assignment is an unlikely explanation of the MDA+IRS contrast with the Routine zone at surveys 2 to 4, when prevalence was lower in every MDA+IRS health area than in every Routine health area (area-level permutation test). Part of the contrast from survey 3 onwards reflects a rise in Routine prevalence that the data do not explain, which occurred in all five Routine health areas (Additional file 1: S8 Table). Season does not explain the rise, because surveys 1 and 3 both followed the low-transmission season. If the rise was unrelated to the campaign, it made the MDA+IRS zone appear further below the Routine zone. With Awasi as a fixed comparator, the contrast was similar, which excludes the changing Routine composition but not the rise, which Awasi shared.

Regression to the mean is also unlikely, because the MDA+IRS zone started with lower prevalence than the Routine zone. Residual confounding cannot be excluded. Because the models were adjusted for baseline prevalence, such a confounder would have had to change prevalence differently in the two zones after baseline. Both the within-survey OR and the DiD ratio of ORs were below 1 at every survey after baseline, meaning that the contrast does not depend on how the baseline difference is handled.

### IRS alone and adding MDA

The difference between the IRS and Routine zones was not statistically significant at any survey in the area-level permutation test, which does not adjust for the higher baseline prevalence of the IRS zone. After the campaign, the within-survey OR for the IRS zone was not statistically significant, whereas the DiD ratio of ORs was, because the IRS zone retained higher odds than the Routine zone at survey 1 after adjustment. If part of that difference was temporary, regression to the mean would lower the DiD ratio of ORs without any effect of IRS. We therefore give more weight to the within-survey OR. Season is an unlikely explanation of the weakening, because surveys 2 and 4 both followed the high-transmission season. The 2023 ITN distributions may have contributed at survey 5, although adjustment for individual net use changed the IRS ORs little. Chance cannot be excluded, because the IRS intervals at surveys 4 and 5 were wide. Survey 4 took place 4.5 to 8.5 months after IRS round 2, within or before the 6.5 to 11 months of clothianidin activity reported in bioassays with susceptible laboratory strains [14,19,20]. However, in Uganda, mud walls retained the least clothianidin [21]. Resurgence there after the switch to clothianidin also coincided with tolerance in *Anopheles funestus* [21]. In this study, residual activity on mud walls in the IRS zone was lost before survey 4, but not on plastic sheeting in Gengere. In Apac District, Uganda, facility test positivity rose back to pre-spraying levels over the 4 to 18 months after IRS stopped [22]. The IRS-zone sporozoite rate rose in each half-year from January 2023 (8 of 107 by July to November 2024), in the direction of the weakening. The weakening may therefore reflect faster loss of clothianidin activity on mud walls than in the published laboratory bioassays, or reduced susceptibility of the vectors in the IRS zone. With one test site per wall type, the two could not be separated.

In the primary model, which adjusts for baseline prevalence, the difference between the MDA+IRS and IRS zones appeared only after the campaign. In the area-level permutation test, which does not, the MDA+IRS zone was below the IRS zone at every survey, but during the campaign by approximately the same margin as at survey 1. MDA may have prolonged the lower prevalence observed with IRS during the campaign. However, the two zones already differed at survey 1 before adjustment for baseline prevalence. An unmeasured confounder as strong as that baseline difference could explain the whole survey-5 difference, and could make the survey-4 difference not statistically significant (Additional file 1: S2 Table, panel F).

### Comparison with published evaluations

The lack of a statistically significant difference between the MDA+IRS and IRS zones during the campaign is consistent with two cluster-randomised trials in moderate-to-high transmission, in which MDA did not reduce prevalence within six months [23,24]. In Zambia, where every arm received insecticide-treated nets and targeted IRS, as both zones did in the Angumu comparison of MDA+IRS with IRS, MDA lowered the odds of infection in children by 72% where baseline prevalence was below 10%, but not in the higher-transmission stratum that corresponds to Angumu [23]. In The Gambia, one round of MDA did not lower prevalence at four to six months [6,24].

In a controlled pre-post evaluation in Katakwi District, a high-transmission area of Uganda, adding co-timed DHA-piperaquine MDA to four rounds of IRS was associated with lower odds of infection, with DiD ORs of 0.28 to 0.66 at surveys during and up to approximately six months after the rounds [25]. In Angumu, the equivalent DiD ratio of ORs was not statistically significant during the campaign and was 0.52 (95% CI 0.33–0.83) at survey 4 and 0.66 (0.46–0.96) at survey 5 (Additional file 1: S2 Table, panel E), within the Katakwi range. In other humanitarian emergency contexts, MDA delivered without IRS was followed within weeks by less self-reported fever in Monrovia, Liberia [26], fewer confirmed malaria cases in Sierra Leone [27], and mixed changes in malaria cases at health facilities in the Central African Republic [28]. In Sierra Leone, where transmission was intense, the fall in cases waned approximately four weeks after each round [27]. Prevalence was not measured more than six months after the last round in the trials [6], in Katakwi [25], or in these emergency evaluations, whereas in Angumu the lower prevalence in the MDA+IRS zone persisted to the last survey, 14 months after the last MDA round.

### Mortality

At surveys 3 and 5, CMR and U5MR in every zone were below the January 2020 levels in Angumu [11], and U5MR was below the rate assumed in the sample size calculation, meaning that only large differences could be detected. In a verbal autopsy study of those deaths reported at survey 1, clinicians attributed 35.9% (95% CI 27.0–45.2; 41 of 115) of under-five deaths to malaria [29], which limits how far a campaign against malaria could lower U5MR. The proportion was similar across zones: 38.2% (13 of 34) in the Routine zone, 41.9% (18 of 43) in the MDA+IRS zone, and 37.5% (9 of 24) in the IRS zone. Mortality was therefore a less sensitive measure of the campaign than malaria prevalence.

The lower CMR in the MDA+IRS zone at survey 3 may partly reflect a difference that predated the campaign, because the survey-3 recall period began 10 weeks before IRS round 1. With person-time restricted from MDA round 1 onwards, to exclude the pre-MDA period, the RR moved towards 1 with a much wider interval (0.78, 0.19–3.17; Additional file 1: S3 Table, panel C). Regression to the mean cannot be excluded, because the MDA+IRS zone started with the highest CMR and had the largest fall. A cause mix favouring malaria does not explain that fall, because households attributed fewer survey-1 deaths to malaria and/or anaemia in the MDA+IRS zone (50.2%) than in the Routine (62.3%) and IRS (58.5%) zones (Additional file 1: S9 Table).

At survey 5, the U5MR point estimate in the MDA+IRS zone was higher than at survey 3, and the CMR and U5MR point estimates were higher than in the IRS zone. The CMR difference from the IRS zone was not statistically significant but close to the threshold (RR 1.69, 0.99–2.88; p=0.057; Additional file 1: S3 Table, panel D). The design-based estimate of this RR was further from the threshold (Additional file 1: S3 Table, panel E). The cause of this higher mortality is not known, but a rebound in malaria deaths is unlikely, because malaria prevalence at survey 5 was lower in the MDA+IRS zone than in the IRS zone.

### Entomological and routine surveillance evidence

In 2023, the sporozoite rate was lower in both intervention zones than in the Routine zone, the direction of the prevalence difference, consistent with shorter mosquito survival after IRS. HBR and EIR did not consistently follow the prevalence ranking, and neither can serve as a zone comparator in this study. Zone HBR in the MDA+IRS zone largely reflects one health area, because 75% to 98% of the *Anopheles* caught there came from Gengere, and without a pre-intervention baseline, zone differences cannot be separated from pre-existing ones. EIR, the product of HBR and the sporozoite rate, inherits both problems. HBR also fell between 2022 and 2024 in 9 of the 10 health areas trapped in both periods, Awasi included, so the fall over time cannot be attributed to the campaign. The health-zone-wide ITN distribution of December 2023, and in Awasi the MSF distribution of August 2023, may have contributed to this fall.

The lower prevalence in the MDA+IRS zone was present both in Gengere and in the other four MDA+IRS health areas, with ORs closer to 1 in Gengere at surveys 3 to 5, but the difference between the two strata in the change from survey 1 was not statistically significant. Besides its swamps, Gengere differed from the other areas in population movement, as reported by programme staff. It has a lake port with boat traffic from Uganda and elsewhere in Ituri. It is also the entry point for displaced people arriving in Angumu and hosts a displacement camp. Gengere had also received IRS in 2021. Because Gengere is the only health area of this kind, its ecology cannot be separated from these differences. The MDA+IRS contrast was therefore not confined to Gengere, but the Gengere estimates cannot be attributed to any one of its features.

The MDA+IRS zone had both the lowest facility TPR and the lowest survey malaria prevalence at every survey after baseline. TPR in the MDA+IRS zone was already below that in the Routine zone before the campaign. It then fell between June and August 2022, during IRS round 1 and MDA rounds 1 and 2, while TPR changed little in the other two zones. From July to November 2023, however, TPR in the MDA+IRS zone was above that in the Routine zone in four of five months, whereas prevalence at survey 4, in October 2023, was approximately half that in the Routine zone. TPR measures positivity among attendees who were tested, so it changes with who attends and who is tested as well as with infection in the population. The agreement in zone ranking at the surveys and the timing of the 2022 fall support the survey finding, but monthly TPR is not a reliable measure of prevalence differences between zones.

### Limitations

The main limitation is that zones were not randomly assigned. MDA+IRS was allocated to the health areas with the highest malaria incidence according to surveillance data, yet malaria prevalence at survey 1 was lowest in this zone. Facility incidence reflects care-seeking and reporting as well as infection, and the 2020 MDA in Lanyi and Ugudo-Zii and IRS in Gengere in 2021 may have lowered prevalence before survey 1. The zones also differed at baseline in mortality, in the proportion displaced, and in receipt of earlier interventions. The models were adjusted for the area-level baseline outcome and for individual displacement status, but the zone contrasts remain associations rather than estimates of the campaign’s effect. E-values indicate that removing the MDA+IRS contrast with the Routine zone at surveys 2 to 5 would require confounding stronger than the baseline imbalance before or after adjustment (Additional file 1: S2 Table, panel F). For the contrast with the IRS zone at surveys 4 and 5, the E-values for the CI limits (1.91 and 1.73) exceeded that of the baseline imbalance after adjustment (1.66) but not before it (2.42), so confounding as strong as the unadjusted baseline difference between those zones could make that contrast no longer statistically significant. However, E-values are based on participant-level ORs and do not address the assignment of zones to 14 health areas. Four of the five Routine health areas later received IRS or MDA, which removed them from the comparison. Future evaluations could randomise the order in which comparison areas receive the intervention, in a stepped-wedge design.

Because follow-up ended at the last survey, 14 months after the last MDA round, the duration of the lower prevalence beyond that point is unknown. At survey 5, the Routine comparator was Awasi alone, the Routine health area with the highest baseline prevalence.

The models allowed for Awasi’s higher starting level, but not for anything that changed malaria in Awasi alone after baseline, so the survey-5 contrasts could be overestimated or underestimated. Spill-over between neighbouring zones and the August 2023 ITN distribution in Awasi would bias the contrasts at surveys 4 and 5 towards the null, although adjustment for net use changed the ORs little.

Recall of deaths over more than a year may have omitted some deaths, with an effect of unknown direction. Overdispersion was not modelled in the mixed models, although design-based estimates, which do not assume Poisson variance, led to the same conclusions (Additional file 1: S3 Table, panel E). The small number of under-five deaths in the Routine zone at survey 5 makes the survey-5 U5MR contrasts imprecise.

No pre-intervention entomological baseline was available, because trapping began approximately four weeks before the first spray round (Additional file 2). Zone differences in the entomological indicators therefore cannot be separated from pre-existing differences. From January 2024, the entomological indicators for the Routine zone were based on Awasi alone, meaning that zone comparisons in 2024 depend on one health area. Mosquitoes were selected for sporozoite testing at random within each trap-week, but with unequal probabilities of selection between trap-weeks, meaning that the zone sporozoite rates may differ from those of the whole catch. Weighting each tested mosquito by the number caught divided by the number tested in its trap-week changed the zone rates by less than 0.4 pp in 11 of 12 zone-half-years. It lowered the MDA+IRS rate from July to December 2023 from 2.4% to approximately 1.3%, which places the MDA+IRS zone further below the Routine zone (Additional file 1: S1 Text).

Care-seeking may have differed between zones, because the IRS zone was allocated partly for its lack of MSF support for routine case management. Differences in RDT supply would change which attendees were tested. Attendance at facilities in another zone, including by displaced people, and HRP2 persisting after MDA in recently treated attendees [30,31] would both make the zone series more similar, towards the null. Abia had no facility data, so the Routine series lacks one of its five health areas. TPR therefore supports the zone ranking from the surveys but does not measure an effect of the campaign.

### Conclusions

In a conflict-affected, holoendemic health zone, co-timed MDA and IRS were associated with lower malaria prevalence to the last survey, 14 months after the last MDA round. This lower prevalence lasted far longer than the protection the drugs give, whereas the association with IRS alone weakened after spraying ended. Where high coverage of both interventions can be achieved, national malaria programmes and humanitarian organisations may consider co-timed MDA and IRS in similar settings, with malaria prevalence monitored in repeated surveys to decide when to repeat the campaign.

## Declarations

### Ethics approval and consent to participate

The MSF Ethics Review Board approved the study protocol (protocol 2216) on 22 March 2022 and its amendments on 26 August 2022, 5 October 2023, and 25 March 2024. The protocol was also approved by the Research Ethics Committee of the University of Kisangani, DRC (UNIKIS/CER/007/2022, March 2022; amendments CER/005/GEAK/2023 and CER/002/GEAK/2024). Household representatives gave verbal informed consent to the questionnaire. Each person tested with an RDT gave written consent, by signature or fingerprint, with an independent witness for illiterate participants. Parents or guardians consented for minors, and children over 10 years of age also gave assent (protocol v3.2, section 6.7.3). Households in the entomological surveillance gave verbal consent to trap placement. Interviewers recorded first names only, and individual information was linked only to a household number during analysis. Electronic databases were password protected and stored on MSF secure servers, with access limited to the study co-investigators and the medical coordinator (protocol v3.2, section 9.3). RDT-positive participants received artemether-lumefantrine free of charge under national protocols.

### Trial registration

Not applicable: this was an observational evaluation of an operational malaria control campaign delivered by the Ministry of Health and MSF; participants were not assigned to interventions by the investigators, and the study was not registered in a trial registry.

### Consent for publication

Not applicable.

### Availability of data and materials

Data used in this study are subject to restrictions arising from participant informed consent and General Data Protection Regulation (GDPR)-aligned privacy protections for identifying and potentially stigmatising health information, and cannot be deposited in a public repository. Anonymised individual-level data are available to qualified researchers upon request to the Epicentre Data Sharing Committee (https://epicentre.msf.org/en/our-projects/study-data-access-request), which reviews all requests independently of the study authors in accordance with Epicentre’s Data Sharing Policy (https://epicentre.msf.org/sites/default/files/2020-02/Epicentre-Data%20Sharing%20Policy-EN-2020.pdf).

Analysis code is available at github.com/jpolonsky/malaria-angumu-2024, archived at Zenodo (https://doi.org/10.5281/zenodo.24021582).

### Authors’ contributions

LG, IC, and EG designed the study and developed the protocol. JP, LG, and SM conducted the data analysis. LG coordinated field data collection. AD designed and supervised the entomological surveillance, provided the cleaned entomological dataset, and reviewed the entomological analysis. HJU provided technical and institutional support. MASM, MM, YK, JPO, PN, ES, and AI coordinated operational aspects of the study. JP and LG drafted the manuscript. All authors contributed to data interpretation and reviewed and approved the final manuscript. JP and LG accessed and verified the underlying data. JP and LG had full access to all data and accept responsibility for the integrity of the data and the accuracy of the analysis.

## Supporting information

Additional file 3: STROBE checklist

Additional file 2: S5 Table (entomology workbook)

## Data Availability

Data used in this study are subject to restrictions arising from participant informed consent and General Data Protection Regulation (GDPR)-aligned privacy protections for identifying and potentially stigmatising health information, and cannot be deposited in a public repository. Anonymised individual-level data are available to qualified researchers upon request to the Epicentre Data Sharing Committee (https://epicentre.msf.org/en/our-projects/study-data-access-request), which reviews all requests independently of the study authors in accordance with Epicentre's Data Sharing Policy. Analysis code is available at https://github.com/jpolonsky/malaria-angumu-2024, archived at Zenodo (https://doi.org/10.5281/zenodo.24021582).

https://github.com/jpolonsky/malaria-angumu-2024

## Acknowledgements

We thank the survey teams, community leaders, and participants in Angumu health zone, and Lisa Ebbers, Wageningen University & Research, for the laboratory and species identification work and the entomological analysis in her MSc thesis [15]. We remember Professor Eric Sompwe Mukomena, Programme National de Lutte contre le Paludisme, Kinshasa, and head of the tropical medicine unit, University of Lubumbashi, who provided technical and institutional support to this study and sadly passed away before its publication.

## List of abbreviations

CDC: Centers for Disease Control and Prevention
CI: confidence interval
CMR: crude mortality rate
DHA-piperaquine: dihydroartemisinin-piperaquine
DiD: difference-in-differences
DRC: Democratic Republic of the Congo
EIR: entomological inoculation rate
ELISA: enzyme-linked immunosorbent assay
HBR: human biting rate
HRP2: histidine-rich protein 2
IRS: indoor residual spraying
ITN: insecticide-treated net
MDA: mass drug administration
MSF: Médecins Sans Frontières
OR: odds ratio
PNLP: Programme National de Lutte contre le Paludisme
pp: percentage points
RDT: rapid diagnostic test
RR: rate ratio
STROBE: Strengthening the Reporting of Observational Studies in Epidemiology
TPR: test positivity rate
U5MR: under-five mortality rate
WHO: World Health Organization

