## Additional file 1: supplementary methods, tables and figures for "Malaria prevalence after mass drug administration and indoor residual spraying in a conflict-affected setting, Democratic Republic of the Congo: a non-randomised repeated cross-sectional study, 2022–2024"

##### Contents


- S1 Text. Supplementary methods: setting and interventions; surveys; prevalence and mortality models; sensitivity and additional analyses; entomological and facility surveillance; software.
- S1 Table. Characteristics of survey participants by zone, survey 1, March and April 2022.
- S2 Table. Sensitivity and additional analyses of malaria prevalence (panels A to H).
- S3 Table. Sensitivity and additional analyses of mortality (panels A to F).
- S4 Table. Clusters and households in the zone comparison, by zone and survey.
- S6 Table. Characteristics of the households and participants enrolled, by survey.
- S7 Table. Coverage of MDA and IRS, by round and zone.
- S8 Table. Malaria prevalence by health area and survey.
- S9 Table. Household-reported cause of death, by zone, survey, and age group.
- S10 Table. Monthly facility test positivity rate by zone, October 2021 to October 2024.
- S11 Table. Bed-net use and ownership by zone, surveys 1, 3, and 5.
- S12 Table. Mortality of *Anopheles* in cone bioassays on walls sprayed in IRS round 2, by wall type and visit.
- S1 Figure. Monthly facility test positivity rate, survey malaria prevalence, and facility case counts, by zone.
- S2 Figure. Crude U5MR and CMR by zone and survey.
- S3 Figure. Monthly entomological indicators by zone.
- S4 Figure. Malaria prevalence by health area and survey.
- S5 Figure. Entomological indicators by 2-month period and zone.


S5 Table is Additional file 2 (workbook). The STROBE checklist is Additional file 3.

### S1 Text

#### 1. Setting, interventions, and timing

##### *1.1 Earlier interventions*

The 2020 mass drug administration (MDA) campaign delivered three rounds from September 2020 to January 2021 [1]. It covered Lanyi and Ugudo-Zii, now in the MDA+IRS zone, and Anzika (since divided into Anzika and Pakala) and Panyadong, now in the IRS zone. Indoor residual spraying (IRS) with pirimiphos-methyl (Actellic 300CS) was delivered in 2021 in Abia (Routine zone) and Gengere (MDA+IRS zone) (study protocol, version 3.2, September 2023, unpublished).

##### *1.2 Interventions and survey timing*

The MDA+IRS zone received four MDA rounds, delivered door to door by community health workers. Rounds 1 and 2 used pyronaridine-artesunate, from 11 to 24 July and from 15 to 21 August 2022, and rounds 3 and 4 used dihydroartemisinin-piperaquine (DHA-piperaquine), in January and February 2023. The MDA+IRS and IRS zones received IRS with SumiShield 50WG (50% clothianidin; Sumitomo Chemical, Tokyo, Japan) at 300 mg of active ingredient per m² [2]. Round 1 ran from 27 June to 10 November 2022, and round 2 from 2 February to 31 May 2023. Table S1-1 gives the time from these rounds to the first and last interview of surveys 3 to 5, in days divided by 30.44. The day on which MDA round 4 ended was not recorded, and 28 February 2023 was assumed.

**Table S1-1. Months from the last MDA round and from IRS round 2 to the first and last interview of surveys 3 to 5, MDA+IRS and IRS zones, Angumu health zone, 2023–2024**

| Survey | Months after the end of MDA round 4 | Months after the end of IRS round 2 | Months after the start of IRS round 2 |
|---|---|---|---|
| 3 | 1.9 to 2.5 | During round 2 | 2.8 to 3.4 |
| 4 | 7.6 to 7.7 | 4.5 to 4.7 | 8.4 to 8.6 |
| 5 | 13.6 to 14.2 | 10.6 to 11.1 | 14.5 to 15.0 |

IRS round 2: 2 February to 31 May 2023. MDA round 4: MDA+IRS zone only; end taken as 28 February 2023.

*1.3 Basis of the coverage estimates*

Coverage of MDA rounds 1 and 2 and IRS round 1 was measured at survey 2, and that of the later rounds at survey 3 (S7 Table). MDA coverage was the proportion of household members listed who had received a round, by document or recall. IRS coverage was based on the household's report that its dwelling had been sprayed, among responding households for round 1 and among household members for round 2.

*1.4 Gengere*

Gengere accounted for 27% of the MDA+IRS zone population (25,172 of 92,869), according to the 2022 health-area estimates of the Bureau Central de la Zone de Santé d'Angumu. It was also sprayed in 2021. Its separation from the other four MDA+IRS areas (S2 Table, panel H; S3 Table, panel F; Additional file 2: S5 Table, sheet B) was defined after the mosquito catch by health area had been examined.

*1.5 Crossover health areas*

Crossover areas were excluded from the zone comparison rather than merged into an intervention zone, because none received the package of the zone it would have joined for a comparable period. Abia and Cawa received IRS one to two months before survey 4, Ara and Linga received MDA without IRS, and Cawa received MDA four months after IRS.

#### 2. Surveys

*2.1 Sample size*

Sample size was calculated for each survey in ENA for SMART 2020 and Stata 16. Survey 1 required 865 households in each of two strata to detect a 1.7-fold ratio in the under-five mortality rate (U5MR) between health areas with and without the 2020 MDA. This calculation assumed 2.6 and 1.5 deaths per 10,000 children per day, 80% power, and a one-sided log-rank test. It also required 140 households per health area to estimate malaria prevalence to within ±5 percentage points (prevalence 50%, design effect 1.5, 10% non-response). The 2,135 households comprised 175 in each of the five areas that had received the 2020 MDA and 140 in each of the other nine.

Survey 2 was sized at 210 households per zone, to detect a relative risk of 1.5 in reported morbidity between the MDA+IRS and IRS zones with 80% power. Survey 4 was also sized at 210 households per zone, to detect a relative risk of 1.5 in malaria prevalence between the MDA+IRS and Routine zones. Survey 3 required approximately 1,040 households per zone to detect a two-fold U5MR difference between the MDA+IRS and Routine zones (1.0 death per 10,000 children per day, 270-day recall). A U5MR ratio of 1.5 between the MDA+IRS and IRS zones would have required 2,889 households per zone and was not a design target. Survey 5 used the survey-1 assumptions, with 865 households per zone.

*2.2 Sampling*

The sampling frame, used at every survey, comprised rooftops on an Airbus satellite image of February 2022, geocoded by the Médecins Sans Frontières (MSF) GIS Centre. A cluster was one randomly selected rooftop and its four nearest neighbours, and all five households were interviewed. Clusters were drawn within health areas at survey 1 and within zones at surveys 2 to 5, and were identified by survey and cluster code. At surveys 3 and 5, malaria testing was restricted to a random subset of households, but approximately 93% of clusters contributed one tested household, too few to estimate a second-stage variance component. Variance was therefore estimated as single-stage, with the cluster as primary sampling unit (PSU), at every survey.

*2.3 Household members and displacement status*

A household member was anyone who had lived in the household for at least one month of the recall period, including those who later left or died and a child born and deceased within that month. Members present at the visit and aged 2 months or older were eligible for malaria testing by rapid diagnostic test (RDT). Members who had left their village of origin because of the violence in Djugu and Mahagi territories were classified as displaced, and others as resident.

*2.4 Survey design and weights*

Design-based estimates used the survey package, with the cluster as PSU, zone as stratum, Taylor linearisation, and no finite population correction. The weight of participant *i* in zone *z* was

$$w_i = (N\_z / n\_z) \times (\bar{h} / h_i),$$

where $N\_z$ is the summed 2022 population of the health areas classified to zone *z* at that survey (§1.4), $n\_z$ the number of participants from zone *z*, $h_i$ the number of members of participant *i*'s household, and $\bar{h}$ the mean of $h_i$ over all households. A health area excluded from the zone comparison contributed to no $N\_z$. At survey 1, clusters were allocated by health area. Using health areas in place of zones as strata at survey 1 changed the crude estimates by at most 1.1 percentage points for malaria prevalence and 0.12 deaths per 10,000 person-days for the crude mortality rate (CMR) and U5MR. The order of the zones on each measure was unchanged. The mixed-effects models were unweighted.

*2.5 Crude estimates*

Crude malaria prevalence was estimated from the survey design, with a *t* 95% confidence interval (CI) on the percentage scale with the design degrees of freedom. CMR and U5MR were ratio estimators of deaths over person-time, per 10,000 person-days, with the lower 95% CI limit truncated at zero. Crude odds ratios (ORs) and rate ratios (RRs) were estimated by quasi-binomial and quasi-Poisson regression on zone, respectively, the latter with a log person-time offset.

*2.6 Prevalence surfaces*

The prevalence surfaces in Figure 4 were derived from cluster-level malaria prevalence, shrunk towards the survey mean by a binomial random-intercept model and smoothed with a Gaussian kernel weighted by the number tested. The bandwidth was chosen by cross-validation for each survey, and survey 2, with 138 mapped clusters, was therefore smoothed more coarsely than the other surveys.

#### 3. Prevalence model

*3.1 Specification*

Malaria prevalence was modelled by mixed-effects logistic regression over the five surveys:

malaria ~ zone × survey + age + sex + displacement status + baseline prevalence + (1 | cluster/household)

Malaria was 1 for a positive RDT and 0 for a negative one, and invalid or missing results were excluded. The references were the Routine zone and survey 1, age in years entered linearly (capped at 100 years), and the random intercepts were for the survey-specific cluster and the household within it. The model had no health-area term, because zone is determined by health area, and its 95% CIs therefore reflect variation between clusters and households only. Season was not entered, because it is collinear with survey.

*3.2 Baseline covariates*

Each model included an area-level baseline covariate from survey 1: malaria prevalence (the unweighted proportion of tested participants who were RDT-positive) in the prevalence model, CMR in the CMR model, and U5MR in the U5MR models. The 14 area values were winsorised at their 95th percentile and standardised over the pooled data, and every participant took the value of their health area. A cluster-level covariate was not possible, because clusters were drawn afresh at each survey.

*3.3 Survey-1 contrasts*

Survey-1 ORs and RRs came from the same models without the baseline covariate, which is calculated from the survey-1 outcome, and are reported without p-values.

*3.4 Within-survey contrasts*

Within each survey, each intervention zone was compared with the concurrent Routine zone with emmeans. ORs were evaluated at the mean age and mean baseline covariate, with the categories of sex and displacement status weighted equally and the random effects set to zero. The model-standardised prevalence difference, in percentage points (pp), was estimated from the same fit and is given beside the OR in Table 1, because an OR lies further from 1 than the prevalence ratio when prevalence is high.

*3.5 Ratio of ORs versus survey 1*

The difference-in-differences (DiD) ratio of ORs for zone $z$ at survey $t$, with odds predicted at fixed covariate values, is

$$\mathrm{RoR}(z, t) = [\mathrm{odds}(z, t) / \mathrm{odds}(z, 1)] / [\mathrm{odds}(\mathrm{Routine}, t) / \mathrm{odds}(\mathrm{Routine}, 1)] = \exp(\beta\_z{\times}t),$$

where $\beta\_z{\times}t$ is the zone-by-survey interaction coefficient. The within-survey OR equals $\mathrm{RoR}(z, t) \times \mathrm{OR}(z, 1)$, where $\mathrm{OR}(z, 1)$ is the residual survey-1 OR from the model with the baseline covariate. The ratio of RRs versus survey 1 for CMR was calculated in the same way.

*3.6 MDA+IRS compared with IRS*

Before adjustment for baseline prevalence, the survey-1 OR comparing the MDA+IRS zone with the IRS zone was 0.43 (95% CI 0.33–0.55). Within each survey, the MDA+IRS zone was also compared with the IRS zone as reference, to estimate the difference associated with adding MDA where IRS was also delivered (S2 Table, panel E).

#### 4. Mortality models

*4.1 Outcome and person-time*

The mortality outcome was the death of a household member during the recall period, which began on 1 January 2021, 18 April 2022, and 9 April 2023 at surveys 1, 3, and 5, respectively, and ended at interview. Person-time ran, without a cap, from the latest of recall start, birth, and arrival to the earliest of death, departure, and interview. Members with no person-time were excluded.

##### *4.2 CMR model*

CMR was modelled by mixed-effects Poisson regression over surveys 1, 3, and 5:

deaths ~ zone × survey + age + sex + displacement status + baseline CMR + offset(log person-time) + (1 | cluster/household)

The terms are defined as in §3.1, and RRs were estimated with emmeans at 10,000 person-days. Each mixed-effects CMR model was fitted with every optimiser available to the allFit function of lme4, from default starting values. The non-singular fit with the highest log-likelihood was kept. The optimisers selected for the primary models and for S2 Table, panels A to H, and S3 Table, panels A to F, are fixed in the code release (§8).

##### *4.3 U5MR models*

U5MR was modelled by mixed-effects Poisson regression at surveys 1, 3, and 5 separately, with zone, sex, displacement status, a log person-time offset, and a cluster random intercept. The survey-3 and survey-5 models also included baseline U5MR, and no U5MR model had an age term. A pooled model did not fit reliably on the small number of under-five deaths, and a household random intercept gave a singular fit. No ratio of RRs versus survey 1 was therefore estimated for U5MR.

#### 5. Sensitivity and additional analyses

S2 Table (malaria prevalence) and S3 Table (mortality) report these analyses, with the methods of each panel in its legend. They address the zone difference remaining at survey 1 after adjustment (panel A of each table), the change in the Routine comparator (panel B of each), and the assignment of zones to only 14 health areas (S2 Table, panels C and D). Further panels address unmeasured confounding (S2 F), bed-net use as a possible mediator (S2 G), person-time before the first MDA round (S3 C), the Poisson variance assumption (S3 E), and the contribution of Gengere (S2 H, S3 F). The Awasi-only, health-area random-intercept, permutation, E-value, net-adjusted, design-based, and Gengere-stratum analyses were not prespecified.

#### 6. Entomological surveillance

##### *6.1 Trapping*

Mosquitoes were collected with miniature Centers for Disease Control and Prevention (CDC) light traps, hung approximately 1.5 m above the floor at the foot of a bed in which one or two adult volunteers slept under a net. Households without a net received a long-lasting insecticidal net. A trap-night was one trap in one house for one night. From May to October 2022, each zone had 15 traps: ten in houses selected at random from the rooftop frame (§2.2) and five in purposively selected traditional houses. Traps were set on four consecutive nights each week and moved weekly within four-week cycles. In October 2022, after catches had fallen following IRS, five traps were withdrawn from the Routine zone and four of them redeployed, two to each intervention zone. Data collected from May 2022 to December 2024 were analysed.

##### *6.2 Data cleaning*

Each trap-night was assigned to its zone from its health area and date. Trap-nights on which the trap did not work were excluded from the denominator, but their mosquitoes remained eligible for sporozoite testing. Trap-nights recorded with a date later than their submission were assigned the submission date (135), and those whose dates had been changed to 2025 in the cleaned data were given the dates of an earlier export (47). Duplicate trap-nights (17), repeat records with identical counts (17), traps set outdoors (5), and trap-nights in

Awasi whose dates could not be verified (52) were removed. The cleaning code is in the code release (§8).

*6.3 Identification and sporozoite testing*

*Anopheles* were identified morphologically to species complex, and a subset to species by PCR [3]. A mosquito was counted as tested for sporozoites when a laboratory result was recorded. Circumsporozoite enzyme-linked immunosorbent assay (ELISA) for *Plasmodium falciparum* was performed at the Laboratory of Entomology, Wageningen University and Research, the Netherlands, from January 2023 to November 2024. The 2022 results, from the Wits Research Institute for Malaria, South Africa, were judged unreliable and were not used [3]. Mosquitoes for ELISA were drawn at random from each trap's weekly catch. From January 2023, 79.0% to 100% of the *Anopheles* caught in each health area and half-year were tested outside Gengere, and 23.2% to 98.8% in Gengere.

*6.4 Definitions*

The sporozoite rate was the number of sporozoite-positive *Anopheles* divided by the number tested. The estimated human biting rate (HBR) was the number of *Anopheles* caught on working trap-nights divided by the number of occupants (people who sleep in the house) summed over the same trap-nights. HBR assumed that every *Anopheles* caught was seeking a host. Because CDC light traps catch fewer mosquitoes than human landing catches [4], HBR was used as an index for comparing zones and periods. The estimated entomological inoculation rate (EIR) was HBR × sporozoite rate × 365, divided by 12 to give infective bites per person per month. Density was the number of *Anopheles* per working trap-night in randomly selected houses.

*6.5 Zones, periods, and weights*

Indicators were estimated for four half-years from July 2022 to June 2024 (P1 to P4) and for July to November 2024 (P5), which match the intervals between surveys. May and June 2022 (P0), mostly before IRS round 1, were analysed separately and were not used as a baseline (Additional file 2: S5 Table, sheet E). Trap-nights were assigned to zones as treated by month: a health area left its zone from the month of its first intervention. Each indicator except the pooled sporozoite rate was averaged with population weights (§1.4) over the health areas trapped in at least three calendar months of the period. Most of the 2-month periods specified in the protocol contained too few sporozoite-positive mosquitoes for a stable EIR (median 5 per zone and period from January 2023). Estimates by 2-month period are given in Additional file 2: S5 Table, sheet F, and in S5 Figure.

*6.6 Estimation and intervals*

The pooled sporozoite rate of a zone, with an exact 95% CI, is the primary estimate, and a population-weighted rate is given in Additional file 2: S5 Table. Other 95% CIs were bootstrap percentile intervals from 2,000 resamples of trapping sites within health areas (500 for the monthly series), reported only where at least 90% of resamples gave a valid estimate. Changes over time were ratios of a zone's values in two periods, calculated over the health areas trapped in both, and each intervention zone's change was also expressed relative to that of the Routine zone. No hypothesis tests were applied.

*6.7 Share of mosquitoes tested and totals*

From January 2023 to November 2024, the proportion of *Anopheles* collected that had a recorded laboratory result was 60.3% in the MDA+IRS zone, 89.6% in the Routine zone, and 98.6% in the IRS zone. The zone estimates from July 2022 to November 2024 were based on 17,591 working trap-nights and 20,354 *Anopheles*. In the zone-by-period cells from January 2023, 7,723 mosquitoes had a sporozoite result, of which 283 were positive.

##### *6.8 Model check and sensitivity variants*

As a check on the population-weighted estimates, a negative binomial mixed model of the catch per working trap-night was fitted, with a likelihood-ratio test of a common change over time across the areas of each zone (Additional file 2: S5 Table, sheet D). The eleven sensitivity variants, none prespecified, are listed in Additional file 2: S5 Table, sheet D (sensitivity).

##### *6.9 Cone bioassays after IRS round 2*

The residual efficacy of IRS round 2 was measured monthly from April to December 2023 by World Health Organization (WHO) cone bioassays with local *Anopheles* [5]. One sprayed wall was tested in each of four mud-walled houses in Panyadong (IRS zone) and four plastic-sheeting shelters in the Gengere camp (MDA+IRS zone). Each test used 12 cones, three on each wall, and four control cones on unsprayed surfaces. Mortality was read at 72 hours, because clothianidin acts slowly [2], and corrected by Abbott's formula. A corrected mortality of at least 80% was taken as effective residual activity, and a test with control mortality above 20% as invalid (S12 Table).

##### *6.10 Selection weighting and monthly EIR*

Each tested mosquito was weighted by the number of *Anopheles* caught divided by the number tested in its trap-week (one trap at one site in one calendar week). Zone sporozoite rates were then recalculated by half-year, to assess the effect of the selection of mosquitoes for ELISA. The weighted rate was within 0.4 pp of the pooled rate in 11 of the 12 zone-half-years from January 2023. The exception was the MDA+IRS zone in July to December 2023, when 23.2% of the *Anopheles* caught in Gengere were tested. In that half-year, the weighted rate was 1.3% (bootstrap 95% CI 0.9–2.6), compared with a pooled rate of 2.4% (exact 95% CI 1.6–3.6). Across sensitivity variants of the weights, it was 1.3% to 2.0%.

EIR was also calculated from monthly HBR and sporozoite rates in each health area, averaged over each half-year and then over the zone. This monthly-averaged EIR was within 15% of the reported EIR in 10 of the 12 zone-half-years, with the same order of the three zones in every half-year. Neither analysis was prespecified.

#### 7. Routine facility surveillance and concordance

##### *7.1 Source and indicator*

Facility data came from the dashboard of the national malaria programme (Programme National de Lutte contre le Paludisme, PNLP). The indicator was the monthly RDT positivity ("Positivité TDR") of each health area, positive RDTs divided by RDTs performed, referred to here as the test positivity rate (TPR), from October 2021 to October 2024. Zone TPR was the unweighted mean of the TPRs of its health areas, with zones as treated by month (S10 Table).

##### *7.2 DHIS2 as a sensitivity source*

As a sensitivity source, zone TPR was also calculated from the provincial District Health Information Software 2 (DHIS2) export, as the sum of positive RDTs divided by the sum of suspected cases. DHIS2 and the PNLP dashboard aggregate the same facility reports, meaning that agreement between them checks data processing only.

##### *7.3 Concordance*

Concordance between facility TPR and survey malaria prevalence was assessed over the 15 zone-by-survey pairs, with TPR taken in April 2022, November 2022, April 2023, October 2023, and April 2024. Because the claim tested is ordinal, we counted the surveys at which the same zone was lowest on both measures, and those at which the three zones ranked identically. A tie counted as no agreement. Kendall's coefficient of concordance (W) over ten rankings of the three zones, one per survey and source, was 0.67 ($\chi^2$ 13.4, 2 degrees of freedom), and 0.67 without survey 4. Its p-value is not interpreted, because values from the same source are not independent. With DHIS2 in place of PNLP data, W was 0.49, the same zone was lowest at three surveys, and the full ranking agreed at one. These measures were not prespecified.

#### 8. Software

Analyses used R 4.5.2 with lme4 2.0.6 (mixed models), emmeans 2.0.4 (contrasts, with multivariate *t* adjustment through mvtnorm 1.4-2), survey 4.5 and srvyr 1.3.1 (design-based estimates), glmmTMB 1.1.15.2 (entomological model check), EValue 4.1.4 (E-values), optimx 2025-4.9 (optimisers for allFit), mgcv 1.9-4 (shrinkage of cluster prevalence for the surfaces), and spatstat 3.6-2 (smoothing of the surfaces). The analysis code, with the optimisers selected for the primary models and for S2 Table, panels A to H, and S3 Table, panels A to F, is available at doi:10.5281/zenodo.24021582.


### S Tables

#### Conventions for the S Tables

Zones are as treated: a crossover health area belongs to the Routine zone until its intervention and is excluded from the zone comparison from the following survey. The Routine zone comprised Abia, Ara, Awasi, Cawa, and Linga at surveys 1 to 3, Ara, Awasi, and Linga at survey 4, and Awasi at survey 5. Survey-weighted estimates use the single-stage cluster design and weights of S1 Text §2.4. Months from baseline are calculated from the midpoint of each survey. Within each survey, contrasts sharing a reference zone carry multivariate *t* 95% CIs and p-values, over two comparisons, or three in the Gengere-stratum models. The CI limits used for the E-values are also multivariate *t*. Not adjusted for multiplicity: crude estimates, DiD ratios of ORs and of RRs, ratios between models, and Gengere ratios of DiD ratios. No adjustment was made across surveys or outcomes. Dashes: not applicable. CI, confidence interval; CMR, crude mortality rate; DHA, dihydroartemisinin; DiD, difference-in-differences; EIR, entomological inoculation rate; HBR, human biting rate; IRS, indoor residual spraying; MDA, mass drug administration; OR, odds ratio; pp, percentage points; PNLP, Programme National de Lutte contre le Paludisme; RDT, rapid diagnostic test; RR, rate ratio; TPR, test positivity rate; U5MR, under-five mortality rate.

**S1 Table. Characteristics of survey participants by zone, survey 1, Angumu health zone, Democratic Republic of the Congo, March and April 2022**

| Characteristic | All zones (N = 9,081) | Routine (N = 2,820) | MDA+IRS (N = 3,748) | IRS (N = 2,513) |
|---|---|---|---|---|
| Sex | | | | |
| Female | 4,977 (55) | 1,544 (54) | 2,052 (56) | 1,381 (57) |
| Male | 4,078 (45) | 1,276 (46) | 1,670 (44) | 1,132 (43) |
| Missing | 26 | 0 | 26 | 0 |
| Age (years), median (IQR) | 15 (6–30) | 16 (6–30) | 15 (6–30) | 15 (5–30) |
| Missing | 6 | 2 | 3 | 1 |
| Age group | | | | |
| 0–4 | 2,017 (21) | 615 (20) | 833 (21) | 569 (22) |
| 5–14 | 2,835 (28) | 879 (28) | 1,169 (29) | 787 (27) |
| 15+ | 4,223 (51) | 1,324 (52) | 1,743 (50) | 1,156 (51) |
| Missing | 6 | 2 | 3 | 1 |
| Displacement status | | | | |
| Resident | 7,104 (76) | 2,342 (82) | 2,653 (67) | 2,109 (80) |
| Displaced | 1,964 (24) | 478 (18) | 1,082 (33) | 404 (20) |
| Missing | 13 | 0 | 13 | 0 |
| Household size, median (IQR) | 4 (3–6) | 4 (3–6) | 5 (4–6) | 4 (3–5) |
| Malaria prevalence | 2,961 (37) | 857 (36) | 994 (31) | 1,110 (47) |
| Missing | 1,642 | 561 | 808 | 273 |
| Deaths in recall period | 438 (4.4) | 106 (3.6) | 249 (5.9) | 83 (3.2) |
| Malaria and/or anaemia deaths | 240 (2.4) | 66 (2.2) | 125 (3.0) | 49 (2.0) |

n (%) or median (interquartile range). Percentages are survey-weighted and counts unweighted. Malaria prevalence: among participants with a valid RDT result; 1,642 of the 9,081 participants had none. Deaths: proportion of all household members enrolled. IRS zone: 83 deaths, including one with no person-time, which is excluded from Table 2 (82 deaths) and from S9 Table (48 malaria and/or anaemia deaths, compared with 49 here). Displacement status: resident or displaced (S1 Text §2.3).

**S2 Table. Sensitivity and additional analyses of malaria prevalence, by zone and survey, Angumu health zone, Democratic Republic of the Congo, 2022–2024**

**(A) DiD ratio of ORs versus survey 1, each intervention zone compared with the Routine zone.** DiD ratio of ORs from the zone-by-survey interaction of the primary model (S1 Text §3.5). Survey-1 rows: residual survey-1 OR from the model with the baseline covariate, without p-value. Crude malaria prevalence: survey-weighted.

| Zone | Survey | Months from baseline | Season | Crude malaria prevalence, % (95% CI) | DiD ratio of ORs (95% CI) | p-value |
|---|---|---|---|---|---|---|
| MDA+IRS | 1 | 0 | Low transmission | 31.4 (28.4–34.3) | 0.71 (0.53–0.94) | — |
| MDA+IRS | 2 | 7 | High transmission | 39.2 (33.3–45.1) | 0.41 (0.27–0.63) | <0.001 |
| MDA+IRS | 3 | 13 | Low transmission | 27.0 (23.0–31.0) | 0.31 (0.22–0.44) | <0.001 |
| MDA+IRS | 4 | 18 | High transmission | 31.9 (24.8–39.0) | 0.27 (0.16–0.47) | <0.001 |
| MDA+IRS | 5 | 24 | Low transmission | 24.4 (20.5–28.3) | 0.36 (0.22–0.58) | <0.001 |
| IRS | 1 | 0 | Low transmission | 46.8 (43.8–49.9) | 1.22 (0.93–1.61) | — |
| IRS | 2 | 7 | High transmission | 54.7 (47.5–62.0) | 0.41 (0.26–0.65) | <0.001 |
| IRS | 3 | 13 | Low transmission | 41.4 (37.2–45.6) | 0.30 (0.21–0.42) | <0.001 |
| IRS | 4 | 18 | High transmission | 57.1 (50.6–63.6) | 0.52 (0.30–0.91) | 0.022 |
| IRS | 5 | 24 | Low transmission | 46.9 (42.2–51.6) | 0.54 (0.33–0.86) | 0.011 |

**(B) Awasi as the Routine comparator at every survey.** Primary model refitted with the Routine zone restricted to Awasi at every survey, which holds the comparator constant while the Routine zone fell from five health areas to three and then one. Routine participants from Abia, Ara, Cawa, and Linga were removed at every survey, and the baseline covariate was calculated over all 14 health areas. Survey-1 rows: residual survey-1 ORs, comparable with panel A. N: participants with a valid RDT result.

| Zone | Survey | Months from baseline | Season | N | Crude malaria prevalence, % (95% CI) | OR (95% CI) | p-value |
|---|---|---|---|---|---|---|---|
| Routine | 1 | 0 | Low transmission | 439 | 44.1 (36.7–51.5) | Reference | — |
| MDA+IRS | 1 | 0 | Low transmission | 2,940 | 31.4 (28.4–34.3) | 0.68 (0.44–1.05) | — |
| IRS | 1 | 0 | Low transmission | 2,240 | 46.8 (43.8–49.9) | 1.06 (0.70–1.62) | — |
| Routine | 2 | 7 | High transmission | 199 | 68.5 (57.3–79.6) | Reference | — |
| MDA+IRS | 2 | 7 | High transmission | 874 | 39.2 (33.3–45.1) | 0.22 (0.11–0.45) | <0.001 |
| IRS | 2 | 7 | High transmission | 845 | 54.7 (47.5–62.0) | 0.35 (0.17–0.72) | 0.003 |
| Routine | 3 | 13 | Low transmission | 254 | 62.0 (53.8–70.2) | Reference | — |
| MDA+IRS | 3 | 13 | Low transmission | 934 | 27.0 (23.0–31.0) | 0.20 (0.12–0.33) | <0.001 |
| IRS | 3 | 13 | Low transmission | 952 | 41.4 (37.2–45.6) | 0.31 (0.19–0.49) | <0.001 |
| Routine | 4 | 18 | High transmission | 165 | 62.8 (55.3–70.4) | Reference | — |
| MDA+IRS | 4 | 18 | High transmission | 836 | 31.9 (24.8–39.0) | 0.24 (0.11–0.50) | <0.001 |
| IRS | 4 | 18 | High transmission | 736 | 57.1 (50.6–63.6) | 0.69 (0.33–1.44) | 0.389 |
| Routine | 5 | 24 | Low transmission | 286 | 56.4 (49.2–63.6) | Reference | — |
| MDA+IRS | 5 | 24 | Low transmission | 875 | 24.4 (20.5–28.3) | 0.28 (0.18–0.46) | <0.001 |
| IRS | 5 | 24 | Low transmission | 942 | 46.9 (42.2–51.6) | 0.67 (0.43–1.05) | 0.089 |

**(C) Health-area random intercept.** ORs from the primary model and from the same model with a health-area random intercept, added because zone was assigned to only 14 health areas and cluster-based 95% CIs may be too narrow. Standard deviation of the health-area intercept: 0.14 on the log-odds scale (variance 0.021).

| Zone | Survey | Months from baseline | OR, primary model (95% CI) | OR with health-area random intercept (95% CI) | p-value |
|---|---|---|---|---|---|
| MDA+IRS | 2 | 7 | 0.35 (0.23–0.54) | 0.35 (0.22–0.56) | <0.001 |
| IRS | 2 | 7 | 0.50 (0.32–0.80) | 0.49 (0.29–0.83) | 0.005 |
| MDA+IRS | 3 | 13 | 0.27 (0.20–0.37) | 0.27 (0.18–0.39) | <0.001 |
| IRS | 3 | 13 | 0.36 (0.26–0.50) | 0.37 (0.24–0.55) | <0.001 |
| MDA+IRS | 4 | 18 | 0.24 (0.13–0.42) | 0.23 (0.12–0.42) | <0.001 |
| IRS | 4 | 18 | 0.64 (0.36–1.14) | 0.62 (0.33–1.16) | 0.158 |
| MDA+IRS | 5 | 24 | 0.31 (0.19–0.50) | 0.34 (0.20–0.60) | <0.001 |
| IRS | 5 | 24 | 0.66 (0.41–1.04) | 0.73 (0.42–1.26) | 0.324 |

**(D) Area-level permutation test.** Difference: unweighted mean of area malaria prevalence (proportion of tested participants who were RDT-positive) in the zone minus that in the comparator (pp). p-value: exact, two-sided, over every division of the health areas of the two zones into groups of their original sizes. With Awasi alone at survey 5, the smallest attainable p-value was 0.167 for MDA+IRS and 0.200 for IRS compared with Routine. The test treats zone labels as exchangeable between health areas, which ignores the allocation of MDA+IRS to the health areas with the highest malaria incidence. Distinct labellings: number of distinct assignments of the zone labels among the health areas present. Survey-1 rows: differences only, without a test. No correction for multiple testing.

| Survey | Months from baseline | Zone | Comparator | Areas (zone / comparator) | Mean area prevalence, % (zone / comparator) | Difference (pp) | Every zone area below every comparator area | Distinct labellings | Permutation p-value |
|---|---|---|---|---|---|---|---|---|---|
| 1 | 0 | MDA+IRS | Routine | 5 / 5 | 34.1 / 38.1 | −4.0 | No | — | — |
| 1 | 0 | IRS | Routine | 4 / 5 | 49.2 / 38.1 | 11.1 | No | — | — |
| 1 | 0 | MDA+IRS | IRS | 5 / 4 | 34.1 / 49.2 | −15.1 | No | — | — |

| Survey | Months from baseline | Zone | Comparator | Areas (zone / comparator) | Mean area prevalence, % (zone / comparator) | Difference (pp) | Every zone area below every comparator area | Distinct labellings | Permutation p-value |
|---|---|---|---|---|---|---|---|---|---|
| 2 | 7 | MDA+IRS | Routine | 5 / 5 | 40.3 / 61.8 | −21.5 | Yes | 252 | 0.008 |
| 2 | 7 | IRS | Routine | 4 / 5 | 55.6 / 61.8 | −6.1 | No | 126 | 0.294 |
| 2 | 7 | MDA+IRS | IRS | 5 / 4 | 40.3 / 55.6 | −15.4 | No | 126 | 0.024 |
| 3 | 13 | MDA+IRS | Routine | 5 / 5 | 25.8 / 55.3 | −29.5 | Yes | 252 | 0.008 |
| 3 | 13 | IRS | Routine | 4 / 5 | 42.5 / 55.3 | −12.8 | No | 126 | 0.183 |
| 3 | 13 | MDA+IRS | IRS | 5 / 4 | 25.8 / 42.5 | −16.7 | No | 126 | 0.024 |
| 4 | 18 | MDA+IRS | Routine | 5 / 3 | 31.0 / 66.1 | −35.1 | Yes | 56 | 0.018 |
| 4 | 18 | IRS | Routine | 4 / 3 | 59.3 / 66.1 | −6.8 | No | 35 | 0.514 |
| 4 | 18 | MDA+IRS | IRS | 5 / 4 | 31.0 / 59.3 | −28.3 | Yes | 126 | 0.016 |
| 5 | 24 | MDA+IRS | Routine | 5 / 1 | 26.2 / 54.2 | −28.0 | Yes | 6 | 0.167 |
| 5 | 24 | IRS | Routine | 4 / 1 | 48.9 / 54.2 | −5.3 | No | 5 | 0.800 |
| 5 | 24 | MDA+IRS | IRS | 5 / 4 | 26.2 / 48.9 | −22.7 | No | 126 | 0.024 |

**(E) MDA+IRS zone compared with the IRS zone.** ORs with the IRS zone as reference (S1 Text §3.6). Survey 1: OR from the model without the baseline covariate, without p-value, and residual survey-1 OR from the model with it.

| Survey | Months from baseline | Model | OR, MDA+IRS vs IRS (95% CI) | p-value | DiD ratio of ORs (95% CI) | p-value |
|---|---|---|---|---|---|---|
| 1 | 0 | Without baseline covariate | 0.43 (0.33–0.55) | — | — | — |
| 1 | 0 | Residual, with baseline covariate | 0.71 (0.53–0.94) | — | — | — |
| 2 | 7 | Primary | 0.70 (0.45–1.09) | 0.138 | 1.00 (0.65–1.54) | 0.985 |
| 3 | 13 | Primary | 0.74 (0.52–1.05) | 0.103 | 1.04 (0.72–1.50) | 0.827 |
| 4 | 18 | Primary | 0.37 (0.23–0.60) | <0.001 | 0.52 (0.33–0.83) | 0.006 |
| 5 | 24 | Primary | 0.47 (0.33–0.68) | <0.001 | 0.66 (0.46–0.96) | 0.030 |

**(F) E-values, MDA+IRS zone compared with the Routine zone at surveys 2 to 5 and with the IRS zone at surveys 4 and 5, with survey-1 benchmarks.** E-values [6]; none for the IRS zone compared with Routine, whose 95% CIs at surveys 4 and 5 included 1. Approximate risk ratio: square root of the OR [7], because malaria was common in the reference zones (crude prevalence 46.9% to 62.5% at the surveys concerned) and no single reference prevalence corresponds to an OR conditional on the random effects and covariates. E-value for the estimate: minimum risk ratio that an unmeasured confounder would need with both zone and malaria, conditional on the model covariates, to explain the OR fully. E-value for the CI limit: the same for the multivariate *t* 95% CI limit closer to 1; 1.00 where the 95% CI includes 1. Benchmark rows: pre-intervention zone difference (survey 1) on the E-value scale, before or after adjustment for baseline prevalence; after adjustment, shrunk towards 1 by construction. The MDA+IRS versus IRS benchmark compares the same health areas as at surveys 4 and 5. The MDA+IRS versus Routine benchmark includes all five original Routine areas, whereas the Routine zone comprised three areas at survey 4 and Awasi alone at survey 5. E-values address confounding at the level of the individual; panels C and D address the assignment of zones to 14 health areas. Not shown: CI-limit E-value for a benchmark row. E-values were calculated with the EValue package [8].

| Comparison | Survey | Row | OR (95% CI) | Approximate risk ratio | E-value | E-value for CI limit |
|---|---|---|---|---|---|---|

| Comparison | Survey | Row | OR (95% CI) | Approximate risk ratio | E-value | E-value for CI limit |
|---|---|---|---|---|---|---|
| MDA+IRS vs Routine | 2 | Within-survey OR | 0.35 (0.23–0.54) | 0.60 | 2.75 | 2.05 |
| MDA+IRS vs Routine | 3 | Within-survey OR | 0.27 (0.20–0.37) | 0.52 | 3.27 | 2.68 |
| MDA+IRS vs Routine | 4 | Within-survey OR | 0.24 (0.13–0.42) | 0.49 | 3.52 | 2.47 |
| MDA+IRS vs Routine | 5 | Within-survey OR | 0.31 (0.19–0.50) | 0.55 | 3.01 | 2.18 |
| MDA+IRS vs IRS | 4 | Within-survey OR | 0.37 (0.23–0.60) | 0.61 | 2.67 | 1.91 |
| MDA+IRS vs IRS | 5 | Within-survey OR | 0.47 (0.33–0.68) | 0.69 | 2.28 | 1.73 |
| MDA+IRS vs Routine | 1 | Survey-1 benchmark: after adjustment for baseline prevalence | 0.87 (0.67–1.11) | 0.93 | 1.36 | — |
| MDA+IRS vs Routine | 1 | Survey-1 benchmark: before adjustment for baseline prevalence | 0.75 (0.59–0.97) | 0.87 | 1.57 | — |
| MDA+IRS vs IRS | 1 | Survey-1 benchmark: after adjustment for baseline prevalence | 0.71 (0.53–0.94) | 0.84 | 1.66 | — |
| MDA+IRS vs IRS | 1 | Survey-1 benchmark: before adjustment for baseline prevalence | 0.43 (0.33–0.55) | 0.66 | 2.42 | — |

**(G) Malaria prevalence adjusted for bed-net use.** Bed-net use is a possible mediator of the zone contrasts. Model 1: primary model with area-level net use at survey 1 (proportion of tested participants who slept under any net the night before, winsorised and standardised as the baseline covariate; correlation with baseline prevalence over the 14 areas −0.46), surveys 1 to 5. Its reference is the primary model refitted with the same optimiser selection, and the primary-OR column gives that refitted value. Model 2: individual net use the night before the survey, with a net-by-survey interaction, surveys 1, 3, and 5, beside a reference model refitted on the same 12,777 participants. Net use was not asked at survey 2, and the survey-4 item could not be verified. Each model was fitted with every allFit optimiser and the non-singular fit with the highest log-likelihood kept. Ratio: zone OR in the net-adjusted model divided by that in its reference model. The optimiser spread of a zone log-odds contrast was 0.027 to 0.120 across these models, above a tolerance of 0.02, so ratios within approximately 3% (primary model) to 13% (Model 1) of 1 lie within optimiser variation. Survey-1 rows: residual survey-1 ORs.

| Model | Survey | Months from baseline | Zone vs Routine | OR, reference model | OR, net-adjusted model (95% CI) | p-value | Ratio |
|---|---|---|---|---|---|---|---|
| 1, area-level net use | 1 | 0 | MDA+IRS | 0.88 | 0.91 (0.70–1.16) | 0.596 | 1.03 |
| 1, area-level net use | 1 | 0 | IRS | 1.24 | 1.41 (1.05–1.90) | 0.019 | 1.14 |
| 1, area-level net use | 2 | 7 | MDA+IRS | 0.35 | 0.36 (0.23–0.55) | <0.001 | 1.01 |
| 1, area-level net use | 2 | 7 | IRS | 0.50 | 0.56 (0.35–0.89) | 0.010 | 1.11 |
| 1, area-level net use | 3 | 13 | MDA+IRS | 0.27 | 0.28 (0.20–0.38) | <0.001 | 1.03 |
| 1, area-level net use | 3 | 13 | IRS | 0.36 | 0.41 (0.29–0.58) | <0.001 | 1.13 |
| 1, area-level net use | 4 | 18 | MDA+IRS | 0.23 | 0.25 (0.14–0.44) | <0.001 | 1.09 |
| 1, area-level net use | 4 | 18 | IRS | 0.63 | 0.74 (0.41–1.33) | 0.392 | 1.17 |
| 1, area-level net use | 5 | 24 | MDA+IRS | 0.30 | 0.32 (0.20–0.52) | <0.001 | 1.05 |
| 1, area-level net use | 5 | 24 | IRS | 0.65 | 0.75 (0.46–1.22) | 0.300 | 1.16 |
| 2, individual net use | 1 | 0 | MDA+IRS | 0.92 (0.73–1.16) | 0.94 (0.75–1.19) | 0.793 | 1.02 |
| 2, individual net use | 1 | 0 | IRS | 1.13 (0.87–1.46) | 1.18 (0.91–1.54) | 0.264 | 1.05 |
| 2, individual net use | 3 | 13 | MDA+IR | 0.27 (0.19–0.36) | 0.26 (0.19–0.36) | <0.001 | 0.98 |

| Model | Survey | Months from baseline | Zone vs Routine | OR, reference model | OR, net-adjusted model (95% CI) | p-value | Ratio |
|---|---|---|---|---|---|---|---|
| | | | S | | | | |
| 2, individual net use | 3 | 13 | IRS | 0.32 (0.23–0.44) | 0.30 (0.21–0.41) | <0.001 | 0.94 |
| 2, individual net use | 5 | 24 | MDA+IRS | 0.34 (0.21–0.55) | 0.34 (0.21–0.56) | <0.001 | 1.01 |
| 2, individual net use | 5 | 24 | IRS | 0.63 (0.39–0.99) | 0.65 (0.41–1.04) | 0.074 | 1.04 |

**(H) Gengere as its own stratum.** Primary model with zone replaced by four strata: the Routine zone (reference), the IRS zone, Gengere, and the other four MDA+IRS health areas (Apala, Lanyi, Ugudo-Zii, and Uyandu), because Gengere supplied most of the *Anopheles* caught in the MDA+IRS zone. The strata were defined after Gengere's survey outcomes had been seen. Primary MDA+IRS and Routine estimates: Table 1. Crude malaria prevalence: survey-weighted with zone weights, with Gengere and the other four MDA+IRS areas each estimated from their own clusters. IRS ORs differ from those of the primary model by at most 0.03 on the log scale. The baseline covariate takes one value in Gengere, so the adjusted Gengere ORs use its slope estimated from the other 13 health areas. The DiD ratios of ORs do not depend on that slope. Survey-1 rows: ORs from the model without the baseline covariate, without p-value. Last column: Gengere's DiD ratio of ORs divided by that of the other four MDA+IRS areas; joint Wald test over surveys 2 to 5, $\chi^2$ 9.16, 4 degrees of freedom, p=0.057. N: participants with a valid RDT result. Gengere at survey 4: 9 clusters.

| Zone | Survey | Months from baseline | N | Crude malaria prevalence, % (95% CI) | OR vs Routine (95% CI) | p-value | DiD ratio of ORs (95% CI) | Gengere vs other MDA+IRS areas, ratio of DiD ratios (95% CI) |
|---|---|---|---|---|---|---|---|---|
| MDA+IRS excluding Gengere | 1 | 0 | 2,420 | 30.4 (27.2–33.6) | 0.72 (0.55–0.96) | — | — | — |
| Gengere | 1 | 0 | 520 | 36.1 (28.7–43.5) | 1.01 (0.63–1.62) | — | — | — |
| IRS | 1 | 0 | 2,240 | 46.8 (43.8–49.9) | 1.78 (1.35–2.34) | — | — | — |
| MDA+IRS excluding Gengere | 2 | 7 | 678 | 40.3 (33.3–47.4) | 0.37 (0.23–0.60) | <0.001 | 0.44 (0.28–0.69) | — |
| Gengere | 2 | 7 | 196 | 36.3 (24.5–48.1) | 0.31 (0.16–0.61) | <0.001 | 0.32 (0.16–0.62) | 0.73 (0.37–1.43); p = 0.354 |
| IRS | 2 | 7 | 845 | 54.7 (47.5–62.0) | 0.52 (0.32–0.84) | 0.004 | 0.40 (0.26–0.63) | — |
| MDA+IRS excluding Gengere | 3 | 13 | 771 | 25.1 (20.7–29.5) | 0.24 (0.16–0.34) | <0.001 | 0.28 (0.19–0.41) | — |
| Gengere | 3 | 13 | 163 | 35.3 (25.2–45.4) | 0.43 (0.23–0.79) | 0.003 | 0.43 (0.23–0.81) | 1.55 (0.81–2.97); p = 0.188 |
| IRS | 3 | 13 | 952 | 41.4 (37.2–45.6) | 0.38 (0.27–0.53) | <0.001 | 0.29 (0.21–0.42) | — |
| MDA+IRS excluding Gengere | 4 | 18 | 679 | 27.2 (20.0–34.4) | 0.19 (0.10–0.35) | <0.001 | 0.22 (0.13–0.39) | — |
| Gengere | 4 | 18 | 157 | 48.2 (29.0–67.3) | 0.49 (0.21–1.19) | 0.146 | 0.50 (0.22–1.14) | 2.26 (1.03–4.96); p = 0.041 |
| IRS | 4 | 18 | 736 | 57.1 (50.6–63.6) | 0.65 (0.35–1.19) | 0.221 | 0.51 (0.29–0.88) | — |
| MDA+IRS excluding Gengere | 5 | 24 | 685 | 21.5 (17.4–25.6) | 0.25 (0.15–0.44) | <0.001 | 0.30 (0.18–0.50) | — |
| Gengere | 5 | 24 | 190 | 34.2 (24.7–43.7) | 0.46 (0.23–0.92) | 0.023 | 0.47 (0.24–0.94) | 1.56 (0.83–2.95); p = 0.166 |
| IRS | 5 | 24 | 942 | 46.9 (42.2–51.6) | 0.65 (0.40–1.07) | 0.102 | 0.51 (0.31–0.82) | — |

**S3 Table. Sensitivity and additional analyses of mortality, by zone and survey, Angumu health zone, Democratic Republic of the Congo, 2022–2024**

**(A) DiD ratio of RRs versus survey 1, CMR.** DiD ratio of RRs from the zone-by-survey interaction of the pooled CMR model (S1 Text §3.5, §4.2). Crude rates: survey-weighted, deaths per 10,000 person-days. Deaths and person-days: unweighted.

| Zone | Survey | Months from baseline | Deaths / person-days | Crude rate (95% CI) | DiD ratio of RRs (95% CI) | p-value |
|---|---|---|---|---|---|---|
| MDA+IRS | 3 | 13 | 106 / 1,996,199 | 0.44 (0.34–0.54) | 0.46 (0.29–0.73) | 0.001 |
| IRS | 3 | 13 | 91 / 2,009,404 | 0.37 (0.28–0.46) | 0.82 (0.48–1.39) | 0.459 |
| MDA+IRS | 5 | 24 | 134 / 1,918,935 | 0.61 (0.47–0.75) | 0.68 (0.36–1.27) | 0.223 |
| IRS | 5 | 24 | 50 / 1,736,690 | 0.26 (0.18–0.33) | 0.61 (0.30–1.26) | 0.186 |

**(B) Awasi as the Routine comparator, CMR.** Pooled CMR model refitted with the Routine zone restricted to Awasi at every survey, as in S2 Table, panel B, with the optimiser selection of the primary CMR model. U5MR was not refitted, because under-five deaths were few. Survey-1 RRs: from the model without the baseline covariate, without p-value.

| Zone | Survey | Months from baseline | Deaths / person-days | Crude rate (95% CI) | RR (95% CI) | p-value |
|---|---|---|---|---|---|---|
| Routine | 1 | 0 | 19 / 217,824 | 0.81 (0.40–1.23) | Reference | — |
| MDA+IRS | 1 | 0 | 249 / 1,607,904 | 1.38 (1.15–1.60) | 1.73 (0.88–3.39) | — |
| IRS | 1 | 0 | 82 / 1,088,318 | 0.71 (0.48–0.95) | 0.84 (0.41–1.72) | — |
| Routine | 3 | 13 | 34 / 531,452 | 0.51 (0.34–0.68) | Reference | — |
| MDA+IRS | 3 | 13 | 106 / 1,996,199 | 0.44 (0.34–0.54) | 0.65 (0.36–1.18) | 0.183 |
| IRS | 3 | 13 | 91 / 2,009,404 | 0.37 (0.28–0.46) | 0.75 (0.42–1.32) | 0.396 |
| Routine | 5 | 24 | 28 / 494,480 | 0.49 (0.29–0.69) | Reference | — |
| MDA+IRS | 5 | 24 | 134 / 1,918,935 | 0.61 (0.47–0.75) | 0.96 (0.52–1.79) | 0.984 |
| IRS | 5 | 24 | 50 / 1,736,690 | 0.26 (0.18–0.33) | 0.56 (0.29–1.08) | 0.092 |

**(C) CMR at survey 3 with person-time from 11 July 2022.** The survey-3 recall period began on 18 April 2022, before IRS round 1 (from 27 June) and MDA round 1 (from 11 July). RR from the pooled model, and from single-survey models with the pooled-model covariates over the full recall period and over person-time from 11 July 2022. Deaths, person-days, and crude rate: from 11 July 2022, unweighted. p-value: model with person-time from 11 July 2022. U5MR was not refitted, because under-five deaths at survey 3 numbered 24 to 39 per zone.

| Zone | Survey | Months from baseline | Deaths / person-days | Crude rate | RR, pooled model (95% CI) | RR, single-survey model, full recall period (95% CI) | RR, single-survey model, from 11 July 2022 (95% CI) | p-value |
|---|---|---|---|---|---|---|---|---|
| Routine | 3 | 13 | 93 / 1,477,088 | 0.63 | Reference | Reference | Reference | — |
| MDA+IRS | 3 | 13 | 81 / 1,555,098 | 0.52 | 0.64 (0.41–0.99) | 0.67 (0.35–1.28) | 0.78 (0.19–3.17) | 0.901 |
| IRS | 3 | 13 | 61 / 1,572,428 | 0.39 | 0.75 (0.49–1.13) | 0.73 (0.44–1.23) | 0.64 (0.19–2.09) | 0.629 |

**(D) MDA+IRS zone compared with the IRS zone, U5MR and CMR.** U5MR from the single-survey models and CMR from the pooled model, with the IRS zone as reference. Each RR equals the RR for the MDA+IRS zone compared with Routine divided by that for the IRS zone compared with Routine, from the same model. No ratio of RRs versus survey 1 was estimated.

| Outcome | Survey | Months from baseline | RR, MDA+IRS vs IRS (95% CI) | p-value |
|---|---|---|---|---|
| CMR | 3 | 13 | 0.86 (0.53–1.38) | 0.695 |
| CMR | 5 | 24 | 1.69 (0.99–2.88) | 0.057 |
| U5MR | 3 | 13 | 0.72 (0.32–1.60) | 0.567 |
| U5MR | 5 | 24 | 2.10 (0.93–4.77) | 0.082 |

**(E) Design-based estimates, U5MR and CMR.** Check on the Poisson variance assumption of the mixed models, which did not model overdispersion. Survey-weighted quasi-Poisson regression with the covariates and offset of the corresponding mixed model, the cluster as PSU, survey and zone as strata, the weights of S1 Text §2.4, and Taylor-linearised variance, beside the RR from the mixed model. Ratio: design-based RR divided by mixed-model RR. With every weight set to 1, the standard errors of the log RRs were 0.71 to 0.97 times those of the mixed models across the 20 contrasts, and the RRs 0.96 to 1.07 times the mixed-model RRs. Survey-weighted RRs and DiD ratios of RRs at surveys 3 and 5, apart from survey-5 U5MR, were 0.87 to 1.10 times the mixed-model values. Survey-1 rows: models without the baseline covariate, without p-values. † Survey-5 U5MR: four under-five deaths in the Routine zone.

| Outcome | Survey | Months from baseline | Comparison | RR, mixed model (95% CI) | p-value | RR, design-based (95% CI) | p-value | Ratio |
|---|---|---|---|---|---|---|---|---|
| U5MR | 1 | 0 | MDA+IRS vs Routine | 1.45 (0.85–2.47) | — | 1.56 (0.94–2.58) | — | 1.07 |
| U5MR | 1 | 0 | IRS vs Routine | 1.12 (0.62–2.00) | — | 1.34 (0.65–2.78) | — | 1.20 |
| U5MR | 3 | 13 | MDA+IRS vs Routine | 0.58 (0.27–1.28) | 0.226 | 0.61 (0.33–1.15) | 0.150 | 1.05 |
| U5MR | 3 | 13 | IRS vs Routine | 0.81 (0.39–1.68) | 0.752 | 0.83 (0.44–1.56) | 0.744 | 1.02 |
| U5MR | 3 | 13 | MDA+IRS vs IRS | 0.72 (0.32–1.60) | 0.567 | 0.74 (0.37–1.49) | 0.526 | 1.03 |
| U5MR | 5 † | 24 | MDA+IRS vs Routine | 2.40 (0.66–8.78) | 0.213 | 1.77 (0.52–6.08) | 0.450 | 0.74 |
| U5MR | 5 † | 24 | IRS vs Routine | 1.14 (0.29–4.47) | 0.954 | 0.82 (0.22–3.03) | 0.899 | 0.71 |
| U5MR | 5 † | 24 | MDA+IRS vs IRS | 2.10 (0.93–4.77) | 0.082 | 2.17 (0.86–5.49) | 0.116 | 1.03 |
| CMR | 1 | 0 | MDA+IRS vs Routine | 1.78 (1.25–2.56) | — | 1.68 (1.17–2.40) | — | 0.94 |
| CMR | 1 | 0 | IRS vs Routine | 0.87 (0.56–1.35) | — | 0.88 (0.54–1.43) | — | 1.01 |
| CMR | 3 | 13 | MDA+IRS vs Routine | 0.64 (0.41–0.99) | 0.042 | 0.56 (0.38–0.84) | 0.002 | 0.88 |
| CMR | 3 | 13 | IRS vs Routine | 0.75 (0.49–1.13) | 0.206 | 0.76 (0.52–1.10) | 0.178 | 1.02 |
| CMR | 3 | 13 | MDA+IRS vs IRS | 0.86 (0.53–1.38) | 0.695 | 0.75 (0.48–1.15) | 0.234 | 0.87 |
| CMR | 3 | 13 | DiD ratio of RRs, MDA+IRS vs Routine | 0.46 (0.29–0.73) | 0.001 | 0.50 (0.32–0.77) | 0.002 | 1.08 |
| CMR | 3 | 13 | DiD ratio of RRs, IRS vs Routine | 0.82 (0.48–1.39) | 0.459 | 0.80 (0.47–1.38) | 0.424 | 0.98 |
| CMR | 5 | 24 | MDA+IRS vs Routine | 0.94 (0.50–1.78) | 0.966 | 0.85 (0.47–1.52) | 0.744 | 0.90 |
| CMR | 5 | 24 | IRS vs Routine | 0.56 (0.28–1.10) | 0.103 | 0.58 (0.33–1.02) | 0.062 | 1.03 |
| CMR | 5 | 24 | MDA+IRS vs IRS | 1.69 (0.99–2.88) | 0.057 | 1.47 (0.88–2.44) | 0.171 | 0.87 |
| CMR | 5 | 24 | DiD ratio of RRs, MDA+IRS vs Routine | 0.68 (0.36–1.27) | 0.223 | 0.74 (0.42–1.32) | 0.309 | 1.10 |
| CMR | 5 | 24 | DiD ratio of RRs, IRS vs Routine | 0.61 (0.30–1.26) | 0.186 | 0.61 (0.32–1.19) | 0.148 | 1.00 |

**(F) Gengere as its own stratum, U5MR and CMR.** U5MR from the single-survey models and CMR from the pooled model, with zone replaced by the four strata of S2 Table, panel H, and fitted as in S1 Text §4.2 and §4.3. Primary MDA+IRS and Routine estimates: Table 2. Survey-1 RRs: models without the baseline covariate, without p-value. DiD ratio of RRs: CMR only. Last column: Gengere's DiD ratio of RRs divided by that of the other four MDA+IRS areas; joint Wald test over surveys 3 and 5, $\chi^2$ 3.70, 2 degrees of freedom, p=0.157. Crude rates: survey-weighted; deaths and person-days: unweighted. ‡ Gengere U5MR: 3 under-five deaths at survey 3 and 8 at survey 5.

| Outcome | Zone | Survey | Months from baseline | Deaths / person-days | Crude rate per 10,000 person-days (95% CI) | RR vs Routine (95% CI) | p-value | DiD ratio of RRs (95% CI) | Gengere vs other MDA+IRS areas, ratio of DiD ratios (95% CI) |
|---|---|---|---|---|---|---|---|---|---|
| U5M | MDA+IRS excluding | 1 | 0 | 58 / | 2.12 (1.50– | 1.53 (0.85– | — | — | — |

| Outcome | Zone | Survey | Months from baseline | Deaths / person-days | Crude rate per 10,000 person-days (95% CI) | RR vs Routine (95% CI) | p-value | DiD ratio of RRs (95% CI) | Gengere vs other MDA+IRS areas, ratio of DiD ratios (95% CI) |
|---|---|---|---|---|---|---|---|---|---|
| R | Gengere | | | 274,650 | 2.74) | 2.75) | | | |
| U5MR | Gengere | 1 | 0 | 10 / 63,085 | 1.26 (0.46–2.06) | 1.11 (0.41–3.00) | — | — | — |
| U5MR | IRS | 1 | 0 | 38 / 231,820 | 1.77 (0.83–2.72) | 1.12 (0.60–2.09) | — | — | — |
| U5MR | MDA+IRS excluding Gengere | 3 | 13 | 21 / 292,591 | 0.75 (0.38–1.11) | 0.66 (0.27–1.60) | 0.577 | — | — |
| U5MR | Gengere | 3 | 13 | 3 / 68,685 | 0.39 (0.00–0.84) | 0.35 (0.06–1.91) ‡ | 0.352 | — | — |
| U5MR | IRS | 3 | 13 | 30 / 357,327 | 0.80 (0.45–1.14) | 0.82 (0.37–1.79) | 0.893 | — | — |
| U5MR | MDA+IRS excluding Gengere | 5 | 24 | 35 / 250,340 | 1.41 (0.79–2.04) | 2.85 (0.71–11.38) | 0.172 | — | — |
| U5MR | Gengere | 5 | 24 | 8 / 94,108 | 0.88 (0.19–1.57) | 1.34 (0.25–7.14) ‡ | 0.935 | — | — |
| U5MR | IRS | 5 | 24 | 18 / 315,925 | 0.61 (0.30–0.91) | 1.11 (0.26–4.67) | 0.995 | — | — |
| CMR | MDA+IRS excluding Gengere | 1 | 0 | 219 / 1,294,884 | 1.52 (1.26–1.78) | 2.00 (1.36–2.96) | — | — | — |
| CMR | Gengere | 1 | 0 | 30 / 313,020 | 0.75 (0.43–1.06) | 0.93 (0.46–1.86) | — | — | — |
| CMR | IRS | 1 | 0 | 82 / 1,088,318 | 0.71 (0.48–0.95) | 0.87 (0.54–1.39) | — | — | — |
| CMR | MDA+IRS excluding Gengere | 3 | 13 | 88 / 1,584,706 | 0.47 (0.35–0.59) | 0.69 (0.41–1.16) | 0.245 | 0.43 (0.26–0.70) | — |
| CMR | Gengere | 3 | 13 | 18 / 411,493 | 0.32 (0.13–0.52) | 0.60 (0.27–1.32) | 0.308 | 0.66 (0.28–1.57) | 1.54 (0.65–3.65); p = 0.330 |
| CMR | IRS | 3 | 13 | 91 / 2,009,404 | 0.37 (0.28–0.46) | 0.74 (0.47–1.14) | 0.256 | 0.82 (0.48–1.39) | — |
| CMR | MDA+IRS excluding Gengere | 5 | 24 | 97 / 1,422,058 | 0.61 (0.45–0.76) | 0.96 (0.46–1.98) | 0.998 | 0.59 (0.31–1.13) | — |
| CMR | Gengere | 5 | 24 | 37 / 496,877 | 0.62 (0.34–0.89) | 1.12 (0.51–2.46) | 0.972 | 1.24 (0.52–2.96) | 2.09 (0.99–4.43); p = 0.055 |
| CMR | IRS | 5 | 24 | 50 / 1,736,690 | 0.26 (0.18–0.33) | 0.55 (0.27–1.14) | 0.134 | 0.62 (0.30–1.27) | — |

#### S4 Table. Clusters and households in the zone comparison, by zone and survey, Angumu health zone, Democratic Republic of the Congo, 2022–2024

| Unit | Survey | Months from baseline | Dates | Routine | MDA+IRS | IRS | Total |
|---|---|---|---|---|---|---|---|
| Clusters | 1 | 0 | Mar–Apr 2022 | 145 | 158 | 139 | 442 |
| Clusters | 2 | 7 | Nov–Dec 2022 | 43 | 69 | 42 | 154 |

| Unit | Survey | Months from baseline | Dates | Routine | MDA+IRS | IRS | Total |
|---|---|---|---|---|---|---|---|
| Clusters | 3 | 13 | Apr–May 2023 | 208 | 207 | 207 | 622 |
| Clusters | 4 | 18 | Oct 2023 | 20 | 42 | 42 | 104 |
| Clusters | 5 | 24 | Apr–May 2024 | 52 | 206 | 206 | 464 |
| Households | 1 | 0 | Mar–Apr 2022 | 650 | 731 | 616 | 1,997 |
| Households | 2 | 7 | Nov–Dec 2022 | 207 | 256 | 201 | 664 |
| Households | 3 | 13 | Apr–May 2023 | 1,025 | 1,024 | 1,004 | 3,053 |
| Households | 4 | 18 | Oct 2023 | 101 | 208 | 207 | 516 |
| Households | 5 | 24 | Apr–May 2024 | 259 | 1,015 | 993 | 2,267 |

Counts include only clusters and households in health areas within the zone comparison at that survey.

**S6 Table. Characteristics of the households and participants enrolled, by survey, Angumu health zone, Democratic Republic of the Congo, 2022–2024**

| Survey | Months from baseline | Dates | Households | Individuals | Female (%) | Median age, yrs (IQR) | Children <5 (%) | Displaced (%) | Malaria prevalence (%) | Data collected |
|---|---|---|---|---|---|---|---|---|---|---|
| 1 | 0 | Mar–Apr 2022 | 1,997 | 9,081 | 55 | 13 (5–29) | 22.2 | 22 | 39.8 | RDT, mortality, CoD |
| 2 | 7 | Nov–Dec 2022 | 664 | 2,747 | 56 | 12 (5–27) | 23.8 | 24 | 52.7 | RDT only |
| 3 | 13 | Apr–May 2023 | 3,053 | 15,990 | 54 | 14 (6–27) | 19.4 | 23 | 42.5 | RDT, mortality, CoD |
| 4 | 18 | Oct 2023 | 621 | 2,597 | 54 | 13 (5–28) | 21.3 | 22 | 48.2 | RDT only |
| 5 | 24 | Apr–May 2024 | 3,028 | 15,853 | 53 | 14 (6–28) | 18.6 | 20 | 35.9 | RDT, mortality, CoD |

All enrolled households and participants, including those in crossover areas after their intervention. Percentages and median age: unweighted. Malaria prevalence: unweighted, among participants with a valid RDT result. Data collected: RDT, malaria testing; mortality, retrospective mortality recall; CoD, household-reported cause of death. IQR, interquartile range.

**S7 Table. Coverage of MDA and IRS by round and zone, MDA+IRS and IRS zones, Angumu health zone, Democratic Republic of the Congo, 2022–2023**

| Intervention | Round | Zone | % (95% CI) |
|---|---|---|---|
| MDA (Pyramax) | Round 1 (Jul 2022) | MDA+IRS | 91.2 (87.9–93.8) |
| MDA (Pyramax) | Round 2 (Aug 2022) | MDA+IRS | 89.9 (86.1–92.8) |
| MDA (DHA-piperaquine) | Round 3 (2023) | MDA+IRS | 85.0 (83.2–86.7) |
| MDA (DHA-piperaquine) | Round 4 (2023) | MDA+IRS | 84.8 (82.9–86.5) |
| IRS (SumiShield 50WG) | Round 1 (Jul–Aug 2022) | IRS | 96.4 (92.0–98.4) |
| IRS (SumiShield 50WG) | Round 1 (Jul–Aug 2022) | MDA+IRS | 96.5 (92.6–98.4) |
| IRS (SumiShield 50WG) | Round 2 (Apr 2023) | IRS | 97.5 (96.3–98.3) |
| IRS (SumiShield 50WG) | Round 2 (Apr 2023) | MDA+IRS | 94.5 (92.2–96.1) |

% (95% CI): survey-weighted, with logit-scale 95% CIs. MDA rounds 1 and 2 and IRS round 1: measured at survey 2 (November and December 2022); MDA rounds 3 and 4 and IRS round 2: at survey 3 (April and May 2023). MDA: household members listed who received the round, by document or recall. IRS round 1: households that answered; no answer for 84 of 256 MDA+IRS households (70 in Gengere) and 6 of 201 IRS households. IRS round 2: household members; MDA+IRS zone excluding Apala and Uyandu, where the question was not asked. Round labels: months of delivery (S1 Text §1.2).

**S8 Table. Malaria prevalence by health area and survey, Angumu health zone, Democratic Republic of the Congo, 2022–2024**

| Zone at survey 1 | Health area | Survey 1 | Survey 2 | Survey 3 | Survey 4 | Survey 5 |
|---|---|---|---|---|---|---|
| Routine | Abia | 29.4 (23.5–35.3) | 55.1 (40.9–69.4) | 54.7 (47.0–62.3) | Excluded | Excluded |
| Routine | Ara | 28.8 (21.9–35.6) | 55.3 (42.2–68.5) | 40.7 (26.3–55.1) | 51.1 (39.8–62.3) | Excluded |
| Routine | Awasi | 44.1 (36.8–51.3) | 68.5 (57.8–79.2) | 62.0 (53.9–70.2) | 62.8 (55.6–70.1) | 56.4 (49.2–63.6) |
| Routine | Cawa | 36.0 (28.8–43.2) | 66.6 (53.5–79.7) | 44.1 (35.2–53.0) | Excluded | Excluded |
| Routine | Linga | 41.3 (35.5–47.2) | 62.4 (50.5–74.2) | 71.2 (57.1–85.4) | 78.5 (66.7–90.3) † | Excluded |
| MDA+IRS | Apala | 25.7 (20.0–31.4) | 41.5 (33.2–49.8) | 31.3 (20.9–41.8) | 37.0 (22.8–51.2) | 24.2 (14.9–33.5) |
| MDA+IRS | Gengere | 36.1 (29.1–43.1) | 36.3 (25.4–47.2) | 35.3 (25.6–45.0) | 48.2 (32.4–63.9) | 34.2 (25.0–43.4) |
| MDA+IRS | Lanyi | 20.4 (15.8–25.0) | 48.3 (18.8–77.8) | 22.6 (16.8–28.5) | 41.1 (12.6–69.7) † | 24.8 (14.7–35.0) |
| MDA+IRS | Ugudo-Zii | 35.8 (29.7–41.8) | 48.6 (35.6–61.7) | 22.6 (14.1–31.1) | 21.9 (15.8–28.1) | 19.8 (13.4–26.2) |
| MDA+IRS | Uyandu | 42.0 (35.6–48.3) | 30.1 (21.1–39.1) | 22.7 (14.7–30.7) | 12.1 (8.5–15.7) | 18.6 (10.8–26.4) |
| IRS | Anzika | 39.2 (32.6–45.7) | 56.6 (34.5–78.7) | 33.9 (25.8–42.0) | 58.6 (46.1–71.1) | 32.1 (23.1–41.2) |
| IRS | Pakala | 47.6 (42.4–52.8) | 46.0 (34.8–57.2) | 52.7 (43.3–62.0) | 54.7 (40.7–68.8) | 45.9 (37.5–54.4) |
| IRS | Panyadong | 54.7 (49.1–60.4) | 57.0 (43.0–71.0) | 45.0 (35.5–54.5) | 69.4 (55.8–83.1) | 53.0 (45.4–60.6) |
| IRS | Udongo Abira | 44.5 (39.2–49.8) | 59.5 (46.9–72.1) | 37.5 (31.2–43.8) | 51.2 (41.9–60.5) | 55.4 (45.1–65.8) |

% (95% CI): survey-weighted, with 95% CIs calculated as in Table 1 (S1 Text §2.5). Health areas are grouped by zone at survey 1. Excluded: crossover area outside the zone comparison from that survey. † Fewer than five clusters: Linga (2) and Lanyi (3) at survey 4.

**S9 Table. Household-reported cause of death by zone, survey, and age group, Angumu health zone, Democratic Republic of the Congo, surveys 1, 3, and 5, 2022–2024**

#### Under five

| Under five | S1 Routine | S1 MDA+IRS | S1 IRS | S3 Routine | S3 MDA+IRS | S3 IRS | S5 Routine † | S5 MDA+IRS | S5 IRS |
|---|---|---|---|---|---|---|---|---|---|
| Malaria and/or anaemia | 27/36 (75) | 38/68 (56) | 29/38 (76) | 20/39 (51) | 15/24 (63) | 14/30 (47) | 2/4 (50) | 22/43 (51) | 10/18 (56) |
| Cough/respiratory | 0/36 (0) | 5/68 (7) | 1/38 (3) | 0/39 (0) | 1/24 (4) | 3/30 (10) | 0/4 (0) | 0/43 (0) | 0/18 (0) |
| Diarrhoea/vomiting | 0/36 (0) | 6/68 (9) | 2/38 (5) | 0/39 (0) | 0/24 (0) | 0/30 (0) | 0/4 (0) | 3/43 (7) | 1/18 (6) |
| Measles | 0/36 (0) | 1/68 (1) | 0/38 (0) | 0/39 (0) | 0/24 (0) | 0/30 (0) | 0/4 (0) | 0/43 (0) | 0/18 (0) |
| Malnutrition/oedema | 1/36 (3) | 2/68 (3) | 0/38 (0) | 1/39 (3) | 1/24 (4) | 0/30 (0) | 0/4 (0) | 0/43 (0) | 0/18 (0) |
| Accident/violence | 1/36 (3) | 4/68 (6) | 0/38 (0) | 0/39 (0) | 0/24 (0) | 0/30 (0) | 0/4 (0) | 1/43 (2) | 0/18 (0) |
| Childbirth complication | 6/36 (17) | 9/68 (13) | 3/38 (8) | 5/39 (13) | 1/24 (4) | 4/30 (13) | 1/4 (25) | 8/43 (19) | 4/18 (22) |
| Other | 0/36 (0) | 2/68 (3) | 2/38 (5) | 13/39 (33) | 6/24 (25) | 7/30 (23) | 1/4 (25) | 6/43 (14) | 2/18 (11) |
| Unknown (don't know / not recorded) | 1/36 (3) | 1/68 (1) | 1/38 (3) | 0/39 (0) | 0/24 (0) | 2/30 (7) | 0/4 (0) | 3/43 (7) | 1/18 (6) |

#### All ages

| All ages | S1 Routine | S1 MDA+IRS | S1 IRS | S3 Routine | S3 MDA+IRS | S3 IRS | S5 Routine | S5 MDA+IRS | S5 IRS |
|---|---|---|---|---|---|---|---|---|---|
| Malaria and/or anaemia | 66/106 (62) | 125/249 (50) | 48/82 (59) | 43/124 (35) | 37/106 (35) | 25/91 (27) | 9/28 (32) | 52/134 (39) | 16/50 (32) |
| Cough/respiratory | 6/106 (6) | 23/249 (9) | 6/82 (7) | 11/124 (9) | 9/106 (8) | 7/91 (8) | 1/28 (4) | 9/134 (7) | 4/50 (8) |
| Diarrhoea/vomiting | 4/106 (4) | 20/249 (8) | 8/82 (10) | 7/124 (6) | 7/106 (7) | 1/91 (1) | 0/28 (0) | 8/134 (6) | 2/50 (4) |

| All ages | S1 Routine | S1 MDA+IRS | S1 IRS | S3 Routine | S3 MDA+IRS | S3 IRS | S5 Routine | S5 MDA+IRS | S5 IRS |
|---|---|---|---|---|---|---|---|---|---|
| Measles | 0/106 (0) | 2/249 (1) | 1/82 (1) | 0/124 (0) | 0/106 (0) | 0/91 (0) | 0/28 (0) | 0/134 (0) | 0/50 (0) |
| Malnutrition/oedema | 4/106 (4) | 4/249 (2) | 0/82 (0) | 1/124 (1) | 5/106 (5) | 0/91 (0) | 0/28 (0) | 4/134 (3) | 3/50 (6) |
| Accident/violence | 8/106 (8) | 26/249 (10) | 2/82 (2) | 9/124 (7) | 2/106 (2) | 4/91 (4) | 1/28 (4) | 7/134 (5) | 2/50 (4) |
| Childbirth complication | 7/106 (7) | 18/249 (7) | 7/82 (9) | 7/124 (6) | 5/106 (5) | 5/91 (5) | 1/28 (4) | 9/134 (7) | 4/50 (8) |
| Other | 9/106 (8) | 27/249 (11) | 8/82 (10) | 45/124 (36) | 39/106 (37) | 46/91 (51) | 16/28 (57) | 39/134 (29) | 18/50 (36) |
| Unknown (don't know / not recorded) | 2/106 (2) | 4/249 (2) | 2/82 (2) | 1/124 (1) | 2/106 (2) | 3/91 (3) | 0/28 (0) | 6/134 (4) | 1/50 (2) |

Cause: as reported by the household respondent from 11 predefined categories, which included malaria and/or anaemia, grouped into nine by combining malnutrition with oedema and accident with violence. Unknown: "do not know" or no recorded cause. n/N (%): unweighted, without statistical comparison; N, deaths in the mortality models in that zone, survey, and age group. † Routine zone at survey 5: Awasi alone, 4 under-five deaths.

**S10 Table. Monthly facility test positivity rate by zone, Angumu health zone, Democratic Republic of the Congo, October 2021 to October 2024**

| Month | Routine TPR | MDA+IRS TPR | IRS TPR | Routine n | MDA+IRS n | IRS n |
|---|---|---|---|---|---|---|
| 2021-10 | 90.5 | 75.1 | 78.0 | 4 | 5 | 4 |
| 2021-11 | 80.2 | 64.5 | 70.3 | 4 | 4 | 4 |
| 2021-12 | 88.9 | 72.4 | 68.4 | 4 | 5 | 4 |
| 2022-01 | 87.5 | 70.6 | 60.4 | 4 | 5 | 4 |
| 2022-02 | 86.9 | 80.7 | 74.2 | 4 | 5 | 4 |
| 2022-03 | 89.7 | 77.8 | 68.8 | 4 | 5 | 4 |
| 2022-04 | 88.3 | 73.0 | 69.0 | 4 | 5 | 4 |
| 2022-05 | 82.8 | 82.7 | 88.8 | 4 | 5 | 4 |
| 2022-06 | 88.0 | 79.3 | 70.7 | 4 | 5 | 4 |
| 2022-07 | 78.3 | 64.8 | 73.3 | 4 | 5 | 4 |
| 2022-08 | 81.2 | 48.4 | 75.5 | 4 | 5 | 4 |
| 2022-09 | 86.6 | 46.8 | 72.4 | 4 | 5 | 4 |
| 2022-10 | 71.2 | 55.8 | 73.1 | 4 | 5 | 4 |
| 2022-11 | 74.2 | 59.1 | 80.5 | 4 | 5 | 4 |
| 2022-12 | 72.0 | 56.0 | 74.9 | 4 | 5 | 4 |
| 2023-01 | 77.4 | 48.6 | 72.8 | 4 | 5 | 4 |
| 2023-02 | 73.9 | 60.4 | 61.7 | 4 | 5 | 4 |
| 2023-03 | 79.1 | 55.6 | 62.6 | 4 | 5 | 4 |
| 2023-04 | 79.5 | 58.7 | 77.2 | 4 | 5 | 4 |
| 2023-05 | 67.5 | 57.2 | 70.5 | 4 | 5 | 4 |
| 2023-06 | 56.3 | 53.4 | 68.5 | 4 | 5 | 4 |
| 2023-07 | 59.4 | 61.7 | 70.9 | 4 | 5 | 4 |
| 2023-08 | 60.0 | 63.1 | 79.1 | 3 | 5 | 4 |
| 2023-09 | 59.2 | 72.2 | 77.2 | 3 | 5 | 4 |
| 2023-10 | 64.0 | 63.0 | 73.4 | 3 | 5 | 4 |
| 2023-11 | 68.6 | 71.2 | 66.9 | 3 | 4 | 2 |
| 2023-12 | 65.8 | 63.0 | 75.7 | 3 | 5 | 4 |
| 2024-01 | 43.0 | 60.8 | 65.4 | 1 | 5 | 4 |
| 2024-02 | 48.1 | 53.5 | 70.0 | 1 | 5 | 4 |
| 2024-03 | 77.5 | 53.7 | 69.6 | 1 | 5 | 4 |
| 2024-04 | 86.2 | 45.2 | 58.1 | 1 | 5 | 4 |

| Month | Routine TPR | MDA+IRS TPR | IRS TPR | Routine n | MDA+IRS n | IRS n |
|---|---|---|---|---|---|---|
| 2024-05 | 62.9 | 41.8 | 58.3 | 1 | 5 | 3 |
| 2024-06 | 62.0 | 39.8 | 56.4 | 1 | 5 | 4 |
| 2024-07 | 65.0 | 42.2 | 53.4 | 1 | 5 | 4 |
| 2024-08 | 41.2 | 54.8 | 53.4 | 1 | 5 | 4 |
| 2024-09 | 74.2 | 50.9 | 60.1 | 1 | 5 | 4 |
| 2024-10 | 77.1 | 57.1 | 54.0 | 1 | 5 | 4 |

TPR (%): unweighted mean of health-area TPRs; n, health areas contributing. Crossover areas excluded from the month of their first intervention: Abia and Cawa from August 2023, Ara and Linga from January 2024. Abia: no records. Source: PNLP Superset dashboard.

**S11 Table. Bed-net use and ownership by zone, Angumu health zone, Democratic Republic of the Congo, surveys 1, 3, and 5, 2022–2024**

| Measure | Survey | Months from baseline | Routine, % (95% CI), n | MDA+IRS, % (95% CI), n | IRS, % (95% CI), n |
|---|---|---|---|---|---|
| Slept under any net | 1 | 0 | 44.1 (38.3–49.9), n = 2,484 | 60.0 (54.6–65.4), n = 3,266 | 53.9 (47.4–60.5), n = 2,300 |
| Slept under any net | 3 | 13 | 49.2 (45.1–53.4), n = 4,824 | 44.2 (40.0–48.3), n = 5,085 | 17.4 (14.2–20.7), n = 5,050 |
| Slept under any net | 5 | 24 | 53.7 (43.6–63.8), n = 1,269 | 61.8 (56.9–66.6), n = 4,974 | 68.0 (63.6–72.4), n = 4,718 |
| Household owns a net | 1 | 0 | 52.3 (46.0–58.6), n = 2,484 | 68.3 (63.3–73.3), n = 3,266 | 64.1 (58.0–70.2), n = 2,300 |
| Household owns a net | 3 | 13 | 62.8 (58.8–66.9), n = 4,817 | 53.1 (48.9–57.3), n = 5,072 | 27.3 (23.3–31.4), n = 5,050 |
| Household owns a net | 5 | 24 | 65.4 (54.3–76.4), n = 1,269 | 70.5 (65.8–75.3), n = 4,964 | 78.0 (73.8–82.2), n = 4,718 |

Rows: slept under any net the night before, all current household members; lives in a household owning at least one net. % (95% CI): survey-weighted proportions with zone strata and person weights; n, unweighted. Net use was not asked at survey 2, and survey 4 is not shown (net-use item not verified, ownership not asked). Survey 5: Abia, Ara, Cawa, and Linga excluded.

**S12 Table. Mortality of *Anopheles* in WHO cone bioassays on walls sprayed in IRS round 2, by wall type and visit, Angumu health zone, Democratic Republic of the Congo, April to December 2023**

| Wall | Visit | Date | Days | Exposed | Control | Control mortality, % | Corrected mortality, % | At least 80% |
|---|---|---|---|---|---|---|---|---|
| Mud | 1 | 13 Apr 2023 | 10 | 139/173 | 10/58 | 17.2 | 76.3 † | — |
| Mud | 2 | 2 May 2023 | 29 | 147/159 | 8/58 | 13.8 | 91.2 | Yes |
| Mud | 3 | 31 May 2023 | 58 | 133/177 | 10/57 | 17.5 | 69.9 † | — |
| Mud | 4 | 13 Jul 2023 | 101 | 142/157 | 14/57 | 24.6 | 87.3 | Invalid |
| Mud | 5 | 4 Aug 2023 | 123 | 151/173 | 9/58 | 15.5 | 84.9 | Yes |
| Mud | 6 | 1 Sep 2023 | 151 | 119/140 | 13/48 | 27.1 | 79.4 | Invalid |
| Mud | 7 | 29 Sep 2023 | 179 | 113/143 | 9/46 | 19.6 | 73.9 | No |
| Mud | 8 | 3 Nov 2023 | 214 | 146/178 | 8/59 | 13.6 | 79.2 | No |
| Mud | 9 | 22 Dec 2023 | 268 | 139/172 | 13/58 | 22.4 | 75.3 | Invalid |
| Plastic sheeting | 1 | 14 Apr 2023 | 8 | 156/158 | 9/56 | 16.1 | 98.5 | Yes |
| Plastic sheeting | 2 | 4 May 2023 | 28 | 146/162 | 6/55 | 10.9 | 88.9 | Yes |
| Plastic sheeting | 3 | 5 Jun 2023 | 60 | 125/159 | 5/58 | 8.6 | 76.6 † | — |
| Plastic sheeting | 4 | 17 Jul 2023 | 102 | 131/156 | 9/60 | 15.0 | 81.1 | Yes |
| Plastic sheeting | 5 | 9 Aug 2023 | 125 | 130/148 | 12/59 | 20.3 | 84.7 | Invalid |
| Plastic sheeting | 6 | 15 Sep | 162 | 157/168 | 12/60 | 20.0 | 91.8 | Yes |

| Wall | Visit | Date | Days | Exposed | Control | Control mortality, % | Corrected mortality, % | At least 80% |
|---|---|---|---|---|---|---|---|---|
| | | 2023 | | | | | | |
| Plastic sheeting | 7 | 19 Oct 2023 | 196 | 161/173 | 15/60 | 25.0 | 90.8 | Invalid |
| Plastic sheeting | 8 | 7 Dec 2023 | 245 | 156/171 | 3/57 | 5.3 | 90.7 | Yes |
| Plastic sheeting | 9 | 27 Dec 2023 | 265 | 129/180 | 5/59 | 8.5 | 69.0 | No |

Mud walls: Panyadong, IRS zone; plastic sheeting: Gengere camp, MDA+IRS zone; four walls per site (S1 Text §6.9). Days: from spraying of the wall in round 2. Mortality: Abbott-corrected, at 72 hours. † 24-hour reading, no 72-hour reading recorded; not assessed against the 80% threshold. Invalid: control mortality above 20%. Exposed and control: mosquitoes, dead/tested. WHO, World Health Organization.

### S Figures

**S1 Figure. Monthly facility test positivity rate, survey malaria prevalence, and facility malaria cases by zone, Angumu health zone, Democratic Republic of the Congo, October 2021 to October 2024**

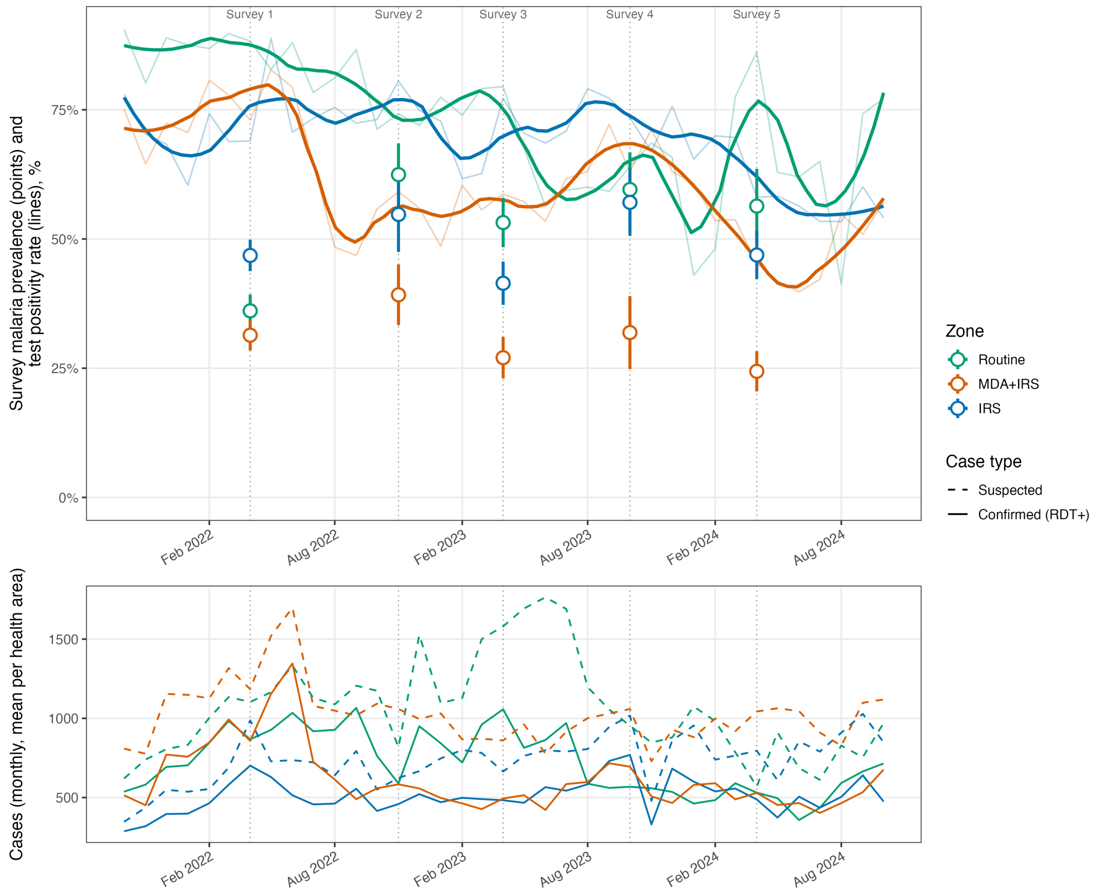


Upper panel: monthly zone TPR (thin lines; PNLP indicator “Positivité TDR”) with a LOESS smooth (thick lines), and survey-weighted malaria prevalence with 95% CI (open points with bars). Lower panel: mean monthly suspected (dashed) and confirmed, RDT-positive (solid) cases per health area. Dotted vertical lines: surveys. Crossover health areas are excluded from the month of their intervention onwards (Abia and Cawa from August 2023, Ara and Linga from January 2024). Abia: no records. Zone TPR is shown without 95% CIs. Values: S10 Table.

**S2 Figure. Crude U5MR and CMR by zone and survey, Angumu health zone, Democratic Republic of the Congo, 2022–2024**

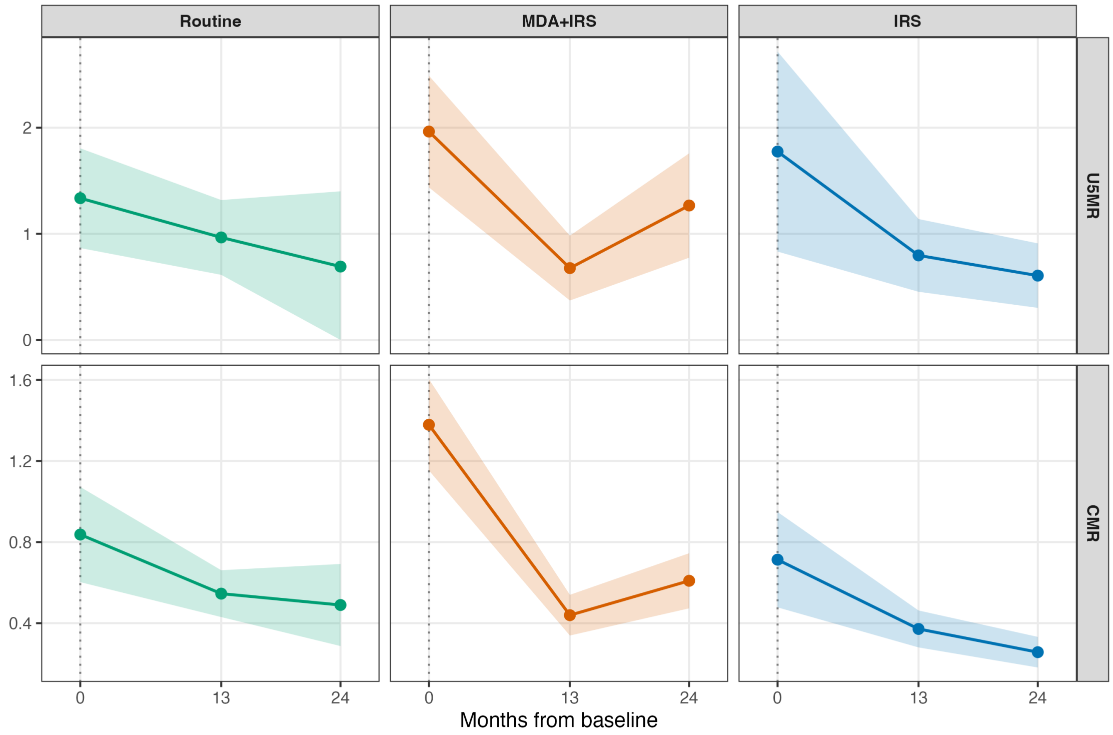


Rows: U5MR and CMR, deaths per 10,000 person-days; columns: zones. Points: survey-weighted rates at 0, 13, and 24 months from baseline; ribbons: 95% CIs. † Routine U5MR at 24 months: lower 95% CI limit truncated at zero (0.69, 95% CI 0.00–1.40). Routine zone at 24 months: Awasi alone.

**S3 Figure. Monthly entomological indicators by zone, Angumu health zone, Democratic Republic of the Congo, May 2022 to November 2024**

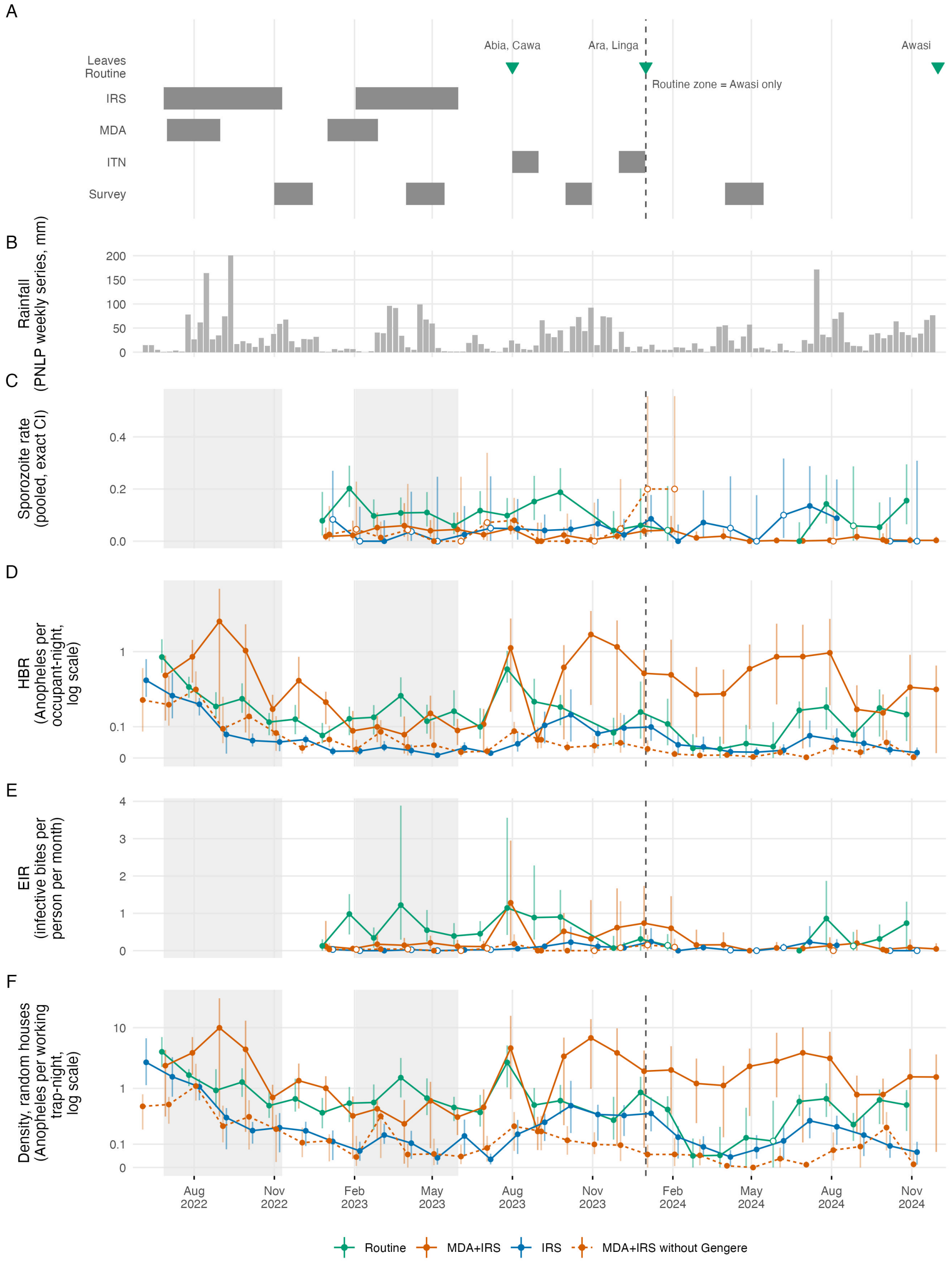


(A) Timing of IRS, MDA, the ITN distributions in Awasi (August 2023) and nationally (December 2023), and surveys 2 to 5, with the month each health area left the Routine zone; dashed vertical line, Routine zone restricted to Awasi from January 2024. (B) Weekly rainfall (PNLP series, mm). (C) Pooled sporozoite rate, exact 95% CIs. (D) HBR, *Anopheles* per occupant-night. (E) EIR, infective bites per person per month. (F) Density, *Anopheles* per working trap-night, randomly selected houses. C and E: from January 2023, months with at least 10 mosquitoes tested. D and F: pseudo-logarithmic axes. Dotted line: MDA+IRS zone without Gengere. Bars in D to F: bootstrap 95% CIs, 500 resamples. Grey bands: IRS rounds. Open symbols: fewer than 30 mosquitoes tested (C, E), or 20 working trap-nights (D), or 20 in randomly selected houses (F). ITN, insecticide-treated net.

**S4 Figure. Malaria prevalence by health area and survey, Angumu health zone, Democratic Republic of the Congo, 2022–2024**

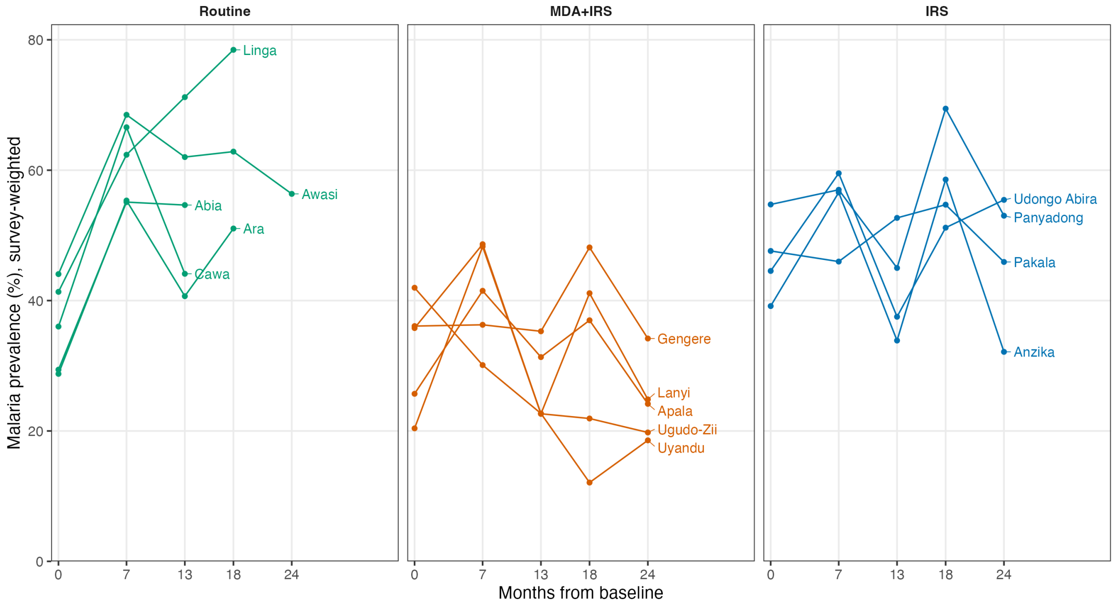


One panel per zone at survey 1; one line per health area, survey-weighted prevalence at 0, 7, 13, 18, and 24 months from baseline. A line stops when the health area left the zone comparison. 95% CIs are not shown; values and 95% CIs: S8 Table.

**S5 Figure. Entomological indicators by 2-month period and zone, Angumu health zone, Democratic Republic of the Congo, May 2022 to November 2024**

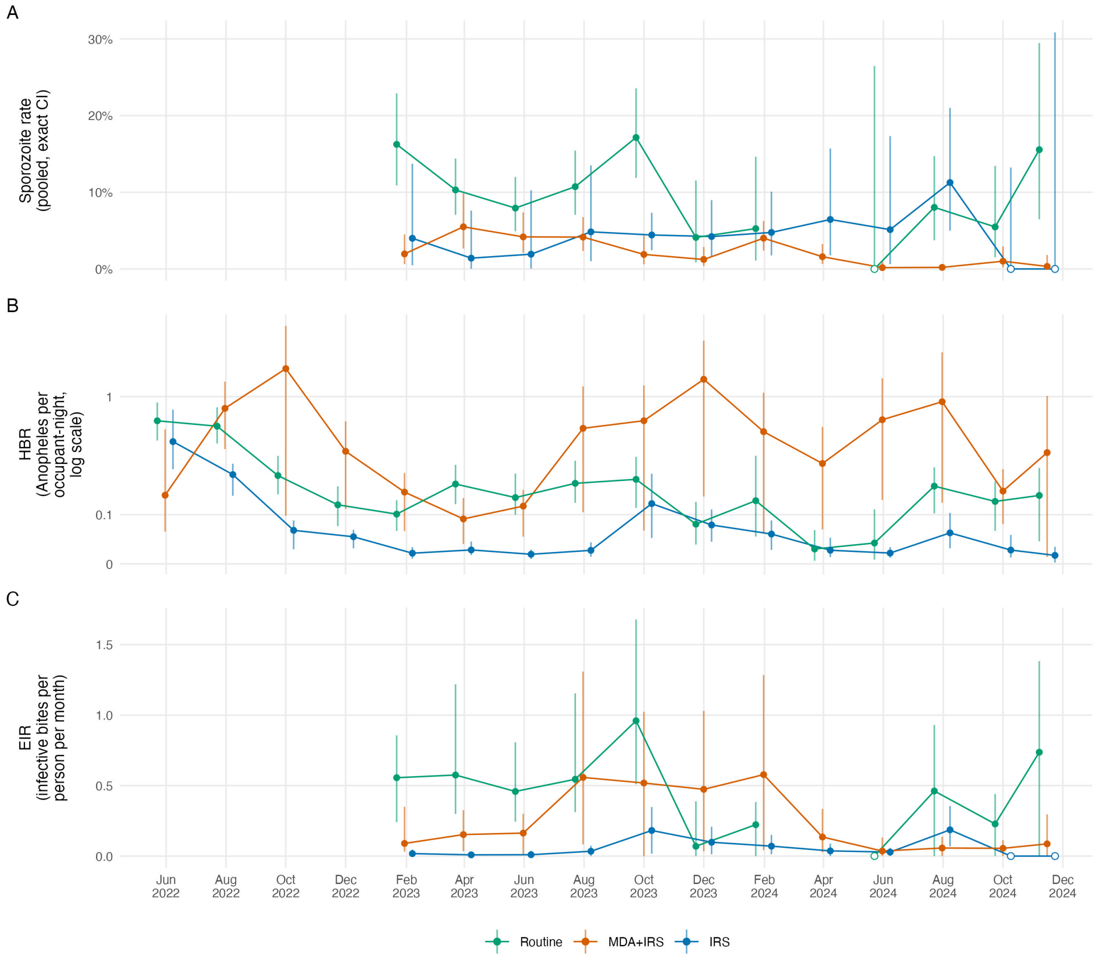

(A) Pooled sporozoite rate, exact 95% CIs, from January 2023. (B) HBR, *Anopheles* per occupant-night, pseudo-logarithmic axis. (C) EIR, infective bites per person per month, from January 2023. Panels A and C: periods with at least 10 mosquitoes tested; open symbols, fewer than 30. Bars in B and C: bootstrap 95% CIs, 2,000 resamples. Points: middle of each 2-month period; November 2024 is a single month. Values: Additional file 2: S5 Table, sheet F.
