## Additional file 3: STROBE checklist for "Malaria prevalence after mass drug administration and indoor residual spraying in a conflict-affected setting, Democratic Republic of the Congo: a non-randomised repeated cross-sectional study, 2022–2024"

### STROBE checklist for cross-sectional studies

Items of the STROBE statement (Strengthening the Reporting of Observational Studies in Epidemiology, cross-sectional studies) and where each is reported. Main-text locations are named by their section headings; Additional file 1 holds S1 Text, the S Tables except S5 Table, and the S Figures; Additional file 2 holds S5 Table.

| Item | Recommendation | Reported in |
|---|---|---|
| **Title and abstract** | | |
| 1 (a) | Indicate the study’s design with a commonly used term in the title or the abstract | Title (“a non-randomised repeated cross-sectional study”); Abstract, Methods |
| 1 (b) | Provide in the abstract an informative and balanced summary of what was done and what was found | Abstract |
| **Introduction** | | |
| 2 | Explain the scientific background and rationale for the investigation being reported | Background, paragraphs 1 to 5 |
| 3 | State specific objectives, including any prespecified hypotheses | Background, final paragraph (objective; primary outcome, malaria prevalence; secondary outcomes, U5MR and CMR) |
| **Methods** | | |
| 4 | Present key elements of study design early in the paper | Methods, Study setting and design; Figure 2 |
| 5 | Describe the setting, locations, and relevant dates, including periods of recruitment, exposure, follow-up, and data collection | Methods, Study setting and design (22 health areas, 14 studied); Interventions (MDA and IRS rounds, crossover dates); Surveys and outcomes (survey dates, recall periods); Entomological surveillance (May 2022 to December 2024); Routine facility surveillance (October 2021 to October 2024); Figures 1 and 2; Additional file 1: S1 Text §1, S6 Table |
| 6 (a) | Give the eligibility criteria, and the sources and methods of selection of participants | Methods, Surveys and outcomes (rooftop-frame cluster sampling; household members aged 2 months or older present at the visit tested; random subset of households tested at surveys 3 and 5); Additional file 1: S1 Text §2.2 and §2.3, S4 Table |
| 7 | Clearly define all outcomes, exposures, predictors, potential confounders, and effect modifiers. Give diagnostic criteria, if applicable | Methods, Interventions (zones and the as-treated classification); Surveys and outcomes (malaria prevalence, U5MR, CMR, displacement status); Statistical analysis (covariates); Entomological surveillance (sporozoite rate, HBR, EIR); Routine facility surveillance (TPR); Additional file 1: S1 Text §2.3, §3.1, §4.1, and §6.4 |
| 8 | For each variable of interest, give sources of data and details of methods of assessment (measurement). Describe comparability of assessment methods if there is more than one group | Methods, Surveys and outcomes (RDT, retrospective mortality recall); Entomological surveillance (CDC light traps, morphological identification, circumsporozoite ELISA); Routine facility surveillance (PNLP facility reports). The same questionnaire, test, and traps were used in every zone. Additional file 1: S1 Text §2, §6.1 to §6.3, and §7.1 |
| 9 | Describe any efforts to address potential sources of bias | Methods, Study setting and design (non-randomised assignment and its basis); Interventions (as-treated classification); Statistical analysis (area-level baseline |

| Item | Recommendation | Reported in |
|---|---|---|
| | | covariate, displacement status, cluster and household random intercepts, sensitivity analyses); Discussion, Limitations; Additional file 1: S1 Text §1.5 and §5 |
| 10 | Explain how the study size was arrived at | Methods, Surveys and outcomes (sample size calculated separately for each survey); Additional file 1: S1 Text §2.1 |
| 11 | Explain how quantitative variables were handled in the analyses. If applicable, describe which groupings were chosen and why | Methods, Statistical analysis (age continuous; displacement status binary; area-level baseline prevalence and mortality as continuous covariates); Additional file 1: S1 Text §3.1, §3.2, and §4.1 (person-time used as recorded) |
| 12 (a) | Describe all statistical methods, including those used to control for confounding | Methods, Statistical analysis; Additional file 1: S1 Text §2.4, §2.5, and §3 to §5 |
| 12 (b) | Describe any methods used to examine subgroups and interactions | Methods, Statistical analysis (zone-by-survey interaction; within-survey contrasts; MDA+IRS compared with IRS); Additional file 1: S1 Text §3.4 to §3.6; S3 Table, panel D |
| 12 (c) | Explain how missing data were addressed | Additional file 1: S1 Text §3.1 (participants with an invalid or missing RDT result excluded) and §4.1 (members with no person-time excluded); S1 Table (missing values by characteristic) |
| 12 (d) | If applicable, describe analytical methods taking account of sampling strategy | Methods, Statistical analysis (survey-weighted estimates with the cluster as the only sampling stage; random intercepts for survey-specific cluster and household within cluster); Additional file 1: S1 Text §2.4 |
| 12 (e) | Describe any sensitivity analyses | Methods, Statistical analysis (Awasi-only comparator, health-area random intercept, area-level permutation test, E-values, net use; for mortality, Awasi-only comparator, person-time from MDA round 1, and design-based estimates); Additional file 1: S1 Text §5, S2 and S3 Tables |
| **Results** | | |
| 13 (a) | Report numbers of individuals at each stage of study, e.g. numbers potentially eligible, examined for eligibility, confirmed eligible, included in the study, completing follow-up, and analysed | Results, Study population and intervention coverage; Additional file 1: S4 Table (clusters and households by zone and survey), S6 Table (households and individuals by survey); Table 1 (participants with a valid RDT result); Table 2 (deaths and person-days) |
| 13 (b) | Give reasons for non-participation at each stage | Households that did not give consent were recorded (survey 1: 26 of 2,023 sampled households, coded as missing consent; survey 3: 15 of 3,054; survey 4: 2 of 621, with 37 missing; survey 5: 21 of 3,028; survey 2 not recorded). Households absent at the visit were replaced in the field by the next nearest household and were not logged |
| 13 (c) | Consider use of a flow diagram | Not included. Figure 2B gives the zone of each health area at each survey, and Additional file 1: S4 and S6 Tables give the cluster, household, and participant counts by survey |
| 14 (a) | Give characteristics of study participants (e.g. demographic, clinical, social) and information on exposures and potential confounders | Results, Study population and intervention coverage (survey-1 paragraph); Additional file 1: S1 and S6 Tables |
| 14 (b) | Indicate number of participants with missing data for each variable of interest | Additional file 1: S1 Table (missing values by characteristic); Table 1 (participants with a valid RDT result) |

| Item | Recommendation | Reported in |
|---|---|---|
| 15 | Report numbers of outcome events or summary measures | Tables 1, 2, and 3; Results, Malaria prevalence, Mortality, and Entomological indicators |
| 16 (a) | Give unadjusted estimates and, if applicable, confounder-adjusted estimates and their precision (e.g. 95% confidence interval). Make clear which confounders were adjusted for and why they were included | Tables 1 and 2 (crude and adjusted estimates with 95% CIs); Methods, Statistical analysis (covariates and the reasons for them); Additional file 1: S1 Text §3 and §4 |
| 16 (b) | Report category boundaries when continuous variables were categorised | Not applicable. Age was analysed as a continuous variable; the age groups of S1 Table are descriptive only |
| 16 (c) | If relevant, consider translating estimates of relative risk into absolute risk for a meaningful time period | Table 1 (model-standardised prevalence difference in percentage points); Table 2 (deaths per 10,000 person-days) |
| 17 | Report other analyses done, e.g. analyses of subgroups and interactions, and sensitivity analyses | Results, Malaria prevalence (Awasi-only comparator, health-area random intercept, permutation test, net use, MDA+IRS compared with IRS, E-values); Mortality (DiD ratio of RRs, MDA+IRS compared with IRS, design-based estimates); Entomological indicators; Routine facility surveillance; Additional file 1: S2, S3, S8, S9, S11, and S12 Tables, S1 Text §5 |
| **Discussion** | | |
| 18 | Summarise key results with reference to study objectives | Discussion, Principal findings; Conclusions |
| 19 | Discuss limitations of the study, taking into account sources of potential bias or imprecision. Discuss both direction and magnitude of any potential bias | Discussion, Limitations |
| 20 | Give a cautious overall interpretation of results considering objectives, limitations, multiplicity of analyses, results from similar studies, and other relevant evidence | Discussion, all subsections; Conclusions |
| 21 | Discuss the generalisability (external validity) of the study results | Discussion, Comparison with published evaluations; Conclusions (“in similar settings”) |
| **Other information** | | |
| 22 | Give the source of funding and the role of the funders for the present study and, if applicable, for the original study on which the present article is based | Declarations, Funding; Authors’ contributions |

CDC, Centers for Disease Control and Prevention; CI, confidence interval; CMR, crude mortality rate; DiD, difference-in-differences; EIR, entomological inoculation rate; ELISA, enzyme-linked immunosorbent assay; HBR, human biting rate; IRS, indoor residual spraying; MDA, mass drug administration; PNLP, Programme National de Lutte contre le Paludisme; RDT, rapid diagnostic test; RR, rate ratio; TPR, test positivity rate; U5MR, under-five mortality rate.
